# Evidence for non-specific effects of live Shingles vaccination against all-cause death and hospitalisation in older adults in England: a population-based cohort study

**DOI:** 10.1101/2025.10.16.25338145

**Authors:** Klara Doherty, Natalie E.R. Beveridge, Laura Bonnett, Valerie Decraene, Caroline Jeffery, Marc Y.R. Henrion, Daniel Hungerford, Neil French

## Abstract

**Background:** Non-specific live vaccine effects have been described in paediatric populations. We investigated whether non-specific effects are observed in older adults with live shingles (*Zostavax®*) immunisation.

**Methods:** A population-based cohort study was performed using health records from the Clinical Practice Research Datalink in England. Shingles vaccine eligible individuals aged 70 years and over who had received pneumococcal immunisation were included. All-cause death, all-cause hospitalisation, and infection-associated hospitalisation rates were compared between live shingles vaccinated and unvaccinated person-time using time-to-event and time-to-subsequent-event Cox regression with Andersen-Gill extension. Live shingles vaccine exposure was included as a time-varying exposure. The models were run on an overlap-weighted pseudo-population to minimise confounding.

**Results:** Between September 2013 and June 2019 314,618 participants aged 70 years and over who had received pneumococcal immunisation prior to study entry were identified, of which 55% had live shingles vaccine exposure. The overlap-weighted pseudo-population consisted of 60021 lives shingles vaccine unexposed and 60245.6 exposed participants. Live shingles vaccine exposure in the overlap-weighted pseudo-population was associated with a reduction in all-cause death (hazard ratio 0.64; 95% confidence interval 0.62, 0.66), a reduction in subsequent all-cause hospitalisation (hazard ratio 0.84 (0.81, 0.88)), and a reduction in first and subsequent infection-associated hospitalisation (hazard ratio for first event 0.81 (0.79, 0.83), and for subsequent events 0.75 (0.73, 0.77)).

**Conclusions:** Receipt of live shingles vaccine associates with lower mortality and morbidity in older adults in England. The potential for causal linkage should be validated in robust prospective studies, with major implications for national immunisation policies.

**Summary:** This cohort study finds live shingles vaccine exposure in older adults associates with reduced morbidity and mortality beyond disease-specific protection, which was not explained by measurable confounding. If validated, this has major implications for vaccine choice in national immunisation policies.

## Background

The live shingles vaccine (*Zostavax®*, Merck) was routinely offered to adults in England aged 70 years and over between 2013 and 2023. It associated with a 64% reduction in herpes zoster disease and 81% reduction in post-herpetic neuralgia in the vaccinated population.^1^ As well as disease-specific protection, live vaccines have been observed to provide wider protection from other infectious pathogens and from malignancies, which has been termed non-specific vaccine effects. In paediatric populations Bacillus Calmette–Guérin (BCG), oral polio, and measles vaccination has been associated with reduced all-cause mortality in low-income settings,^2–4^ and reduced hospitalisation and illness in high-income settings.^5,6^ The precise mechanisms of these observations remain unclear but may include beneficial programming of the innate immune system, as well as cross-reactivity and bystander activation of the adaptive immune system, collectively termed heterologous immune effects.^7^

Whether these non-specific live vaccine effects are observed in older adults has not been assessed. Evidence of non-specific live vaccine effects in older adults would change cost-effectiveness evaluation and have important policy implications. However, older age is associated with immunosenescence which may mean live vaccines do not have the same broad effect.^7^ An association between the live shingles vaccine and protection from dementia and cardiovascular disease has been demonstrated but is considered a result of direct protection from varicella zoster virus effects, rather than a non-specific effect.^8,9^

The aging population in the UK and other settings worldwide is driving a growing demand on healthcare systems.^10^ There is an urgent need to develop cost-effective interventions for older adults to alleviate this health-system burden. The only live vaccine that has been used extensively in the older adult population in the UK is the live shingles vaccine (*Zostavax®*). As a vaccine given to control a chronic established viral infection it is somewhat different to the assessment of the measles and BCG vaccines in children but nevertheless provides an opportunity to look for health benefits beyond the immediate prevention of shingles. We have used a population-based cohort study to explore the effect of live shingles vaccine exposure on all-cause mortality, all-cause hospitalisation, and infection-associated hospitalisation in over 70-year-olds in England.

## Methods

### Study design and data source

We performed a retrospective cohort study of live shingles vaccination using routinely collected healthcare data from the Clinical Practice Research Datalink (CPRD) Aurum database and Health Episode Statistics (HES). CPRD Aurum is a longitudinal database of primary health-care data submitted by participating practices across the UK and is representative of the UK population.^11^ Unless patients opt out, anonymised demographic and medical data is collected monthly and quality checked at individual and practice level to ensure validity of these data. CPRD data is anonymously linked to secondary-care data held by HES and to patient- or practice-level socioeconomic data through the Index of Multiple Deprivation (IMD).^11^ This study followed the STROBE (Strengthening the Reporting of Observational Studies in Epidemiology) reporting guidelines (Supplementary Table 1 for checklist).^12^

**Table 1:** Baseline characteristics of overlap-weighted pseudo-population.

|  |  |  |
| --- | --- | --- |
|  | Live shingles vaccine<br>unexposed<br>n=60021 | Live shingles<br>vaccine exposed<br>N=60245.6 |
| Age (mean, SD) | 71.71 (3.07) | 71.70 (3.07) |
| Female sex (n, %) | 31391.4 (52.3%) | 31685.2 (52.6%) |
| Comorbid (n, %) | 37830.4 (63.0%) | 37016.5 (61.4%) |
| Cardiovascular disease | 8488 (14.1%) | 7611 (12.6%) |
| Respiratory disease | 11227.4 (18.7%) | 10580 (17.6%) |
| Liver disease | 595.8 (1.0%) | 433.4 (0.7%) |
| Diabetes Mellitus | 15116.5 (25.2%) | 14446.6 (24.0%) |
| Solid organ malignancy | 11230.8 (18.7%) | 11468.1 (19.0%) |
| Dementia | 1308.6 (2.2%) | 1016.5 (1.7%) |
| Connective tissue disease | 1142.6 (1.9%) | 1165.1 (1.9%) |
| Peptic ulcer disease | 2471.7 (4.1%) | 2273 (3.8%) |
| Updated Charlson Index of Comorbidity* (mean, SD) | 3.95 (1.28) | 3.89 (1.21) |
| Index Multiple Deprivation quintile** (n, %) |  |  |
| 1 | 16267.7 (27.1%) | 16887.3 (28.0%) |
| 2 | 13780.9 (23.0%) | 14305.4 (23.7%) |
| 3 | 11964 (19.9%) | 11931.4 (19.8%) |
| 4 | 9911.8 (16.5%) | 9772.4 (16.2%) |
| 5 | 8096.6 (13.5%) | 7349.1 (12.2%) |
| Geographical region (n, %) |  |  |
| Northeast | 2816.2 (4.7%) | 2721.8 (4.5%) |
| Northwest | 10597.8 (17.7%) | 10386.2 (17.2%) |
| Yorkshire & The Humber | 2325.8 (3.9%) | 2634.5 (4.4%) |
| East Midlands | 1365 (2.3%) | 1544.1 (2.6%) |
| West Midlands | 11134.4 (18.6%) | 11400.5 (18.9%) |
| East of England | 3694.6 (6.2%) | 3895.7 (6.5%) |
| Southwest | 8802.6 (14.7%) | 8766.9 (14.6%) |
| South Central | 7828.8 (13.0%) | 7638.6 (12.7%) |
| London | 6612.1 (11.0%) | 6247.1 (10.4%) |
| Southeast Coast | 4835.7 (8.1%) | 5002.7 (8.3%) |
| Ever smoked (n, %) | 42840 (71.4%) | 42761.1 (71.0%) |
| Rate of attendance at doctor's practice in two years preceding study entry*** (mean, SD) | 0.20 (0.13) | 0.20 (0.12) |
| Number of influenza vaccinations per year in three years preceding study entry**** (mean, SD) | 0.83 (0.32) | 0.84 (0.31) |
| Varicella-Zoster associated disease in five years preceding study entry (n, %) | 2229.9 (3.7%) | 2310.7 (3.8%) |
| *\*a continuous scale of comorbidity which predict mortality<sup>1</sup>; \*\*n=299 participants with missing index of multiple deprivation, corresponding medical practice's index of multiple deprivation was substituted;*<br>*\*\*\*number of weeks in which participant attended their medical practice in two years preceding year of entry to study / total of 104 weeks; \*\*\*\* number of influenza immunisation events in three years preceding study entry/ 3 giving score between 0 and 3*<br>*SD=standard deviation* |  |  |

### Study setting

Data were obtained from primary-care medical practices in England (the other UK regions do not have CPRD-HES linkage) between 1^st^ September 2013 (introduction of live shingles immunisation) and 30^th^ June 2019.

### Study participants

Eligible participants were individuals aged 70 years or older during the study period (birth year 1933 to 1948). Participants with immunocompromising conditions which would make them ineligible for live vaccination were excluded (Supplementary Table 2). To minimise the impact of inherent differences in health-seeking behaviours between vaccinated and unvaccinated populations we limited participants to those who had demonstrated engagement with the UK vaccination schedule and had been immunised with the pneumococcal polysaccharide vaccine (PPV23) which is offered from 65-years of age.^13,14^ Full exclusion criteria described in Supplementary Materials and illustrated in Supplementary Figure 1.

**Table 2:**
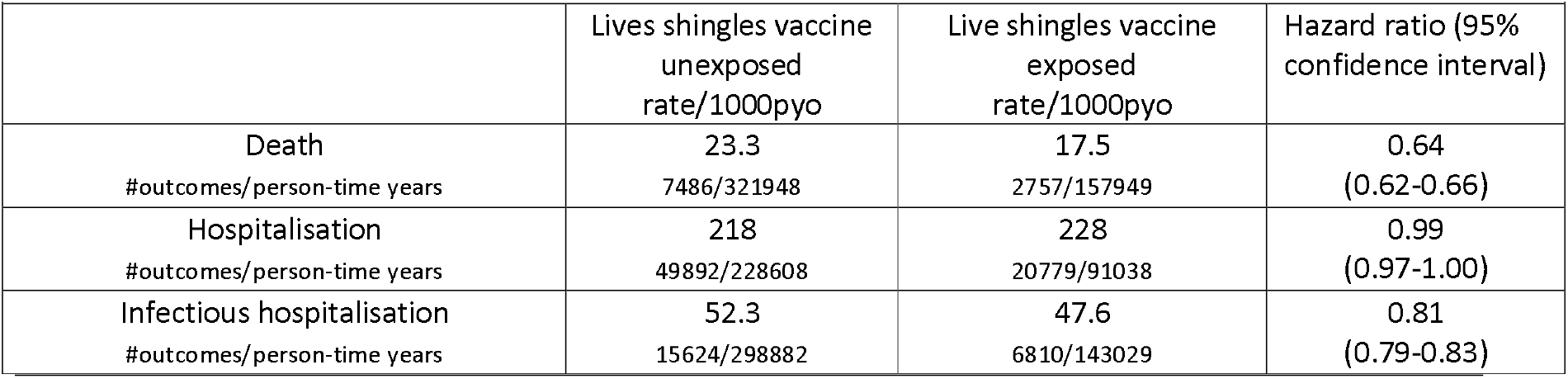
Rate of death and hospitalisation in the overlap-weighted pseudo-population and hazard ratio associated with shingles vaccine exposure.

**Figure 1:**
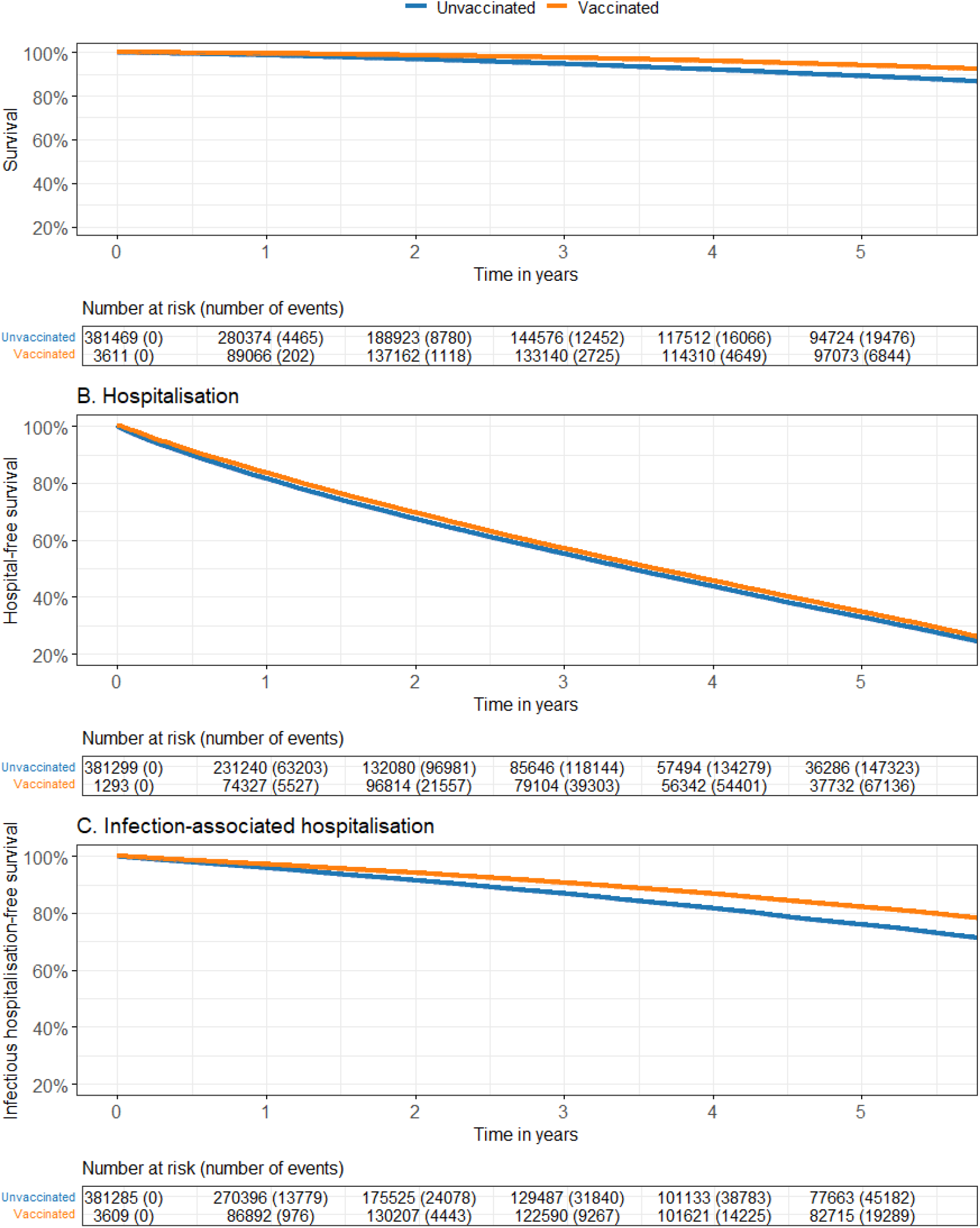
Survival and hospitalisation-free survival in live shingles vaccine exposed and unexposed adults aged 70 years and over. **Survival and hospitalisation-free survival in vaccinated and unvaccinated adults aged ≥70 years**. Survival curves generated using the Simon-Makuch extension of a Kaplan–Meier curve to accommodate time-varying vaccination status. Curves compare time to outcome event during vaccinated and unvaccinated person-time. Time-zero for unvaccinated person-time was study entry and time-zero for vaccinated person-time was date of vaccination. (A) Overall survival (all-cause mortality), (B) All-cause hospitalisation-free survival, (C) Infection-associated hospitalisation–free survival. Numbers at risk at yearly intervals is provided below each panel with cumulative number of events shown in parenthesis.

### Primary exposure: Live shingles vaccination

The primary exposure was receipt of live shingles vaccination, identified from CPRD records (Supplementary Table 3). Vaccination was modelled as a time-varying exposure, with individuals contributing unvaccinated person-time prior to vaccination and vaccinated person-time thereafter.

**Table 3:** Rate of recurrent hospitalisation in the overlap-weighted pseudo-population and instantaneous hazard ratio associated with shingles vaccine exposure.

|  |  |  |  |
| --- | --- | --- | --- |
|  | Lives shingles vaccine<br>unexposed<br>rate/1000pyo | Live shingles vaccine<br>exposed<br>rate/1000pyo | Instantaneous hazard<br>ratio (95% confidence<br>interval) |
| Hospitalisation<br>#outcomes/person-time years | 644<br>255641/397245 | 597<br>122494/205293 | 0.84 (0.81-0.88) |
| Infectious hospitalisation<br>#outcomes/person-time years | 88.5<br>35276/398821 | 77.5<br>15977/206043 | 0.75 (0.73-0.77) |

### Outcome variables: All-cause death, all-cause hospitalisation, and infection-associated hospitalisation

Death during the study period was identified from CPRD. Hospitalisation events were identified from the HES Admitted Patient Care (APC) database. Infection-associated hospitalisation events were identified through the International Classification of Disease and Health Related Problems Revision 10 (ICD-10) coding of primary and secondary discharge diagnoses held by HES using previously defined infectious disease diagnoses (Supplementary Table 4).^15^

### Confounding variables

Baseline variables measured at study entry included: age; sex; comorbid diagnosis (ischaemic heart disease, heart failure, peripheral vascular disease, cerebrovascular disease, dementia, chronic respiratory disease, connective tissue disease, peptic ulcer disease, chronic liver disease, diabetes mellitus, malignancy); the 2011 updated Charlson Comorbidity Index (a continuous scale of comorbidity which predicts mortality in high-income settings)^16^; socioeconomic status (IMD quintile); smoking status; influenza vaccine uptake preceding entry; consultation rate at primary medical practice preceding entry; geographical region of residence; previous varicella zoster disease event; and medical practice. Full description of confounding variables in Supplementary Materials. Baseline variables were measured prior to vaccination and thus were not affected by prior exposure and preserves temporal ordering between exposure and outcome even in the case of time-varying exposure.^17^

### Data collection and measurement

Data used in this study came from routinely collected healthcare data and was not originally intended for use in research. As the data are used for healthcare remuneration, missing vaccine events and hospitalisation were judged to occur rarely. Misclassification of exposure may occur if an event date is entered inaccurately. CPRD applies data quality checks to ensure valid age, sex, death date, and clinical event dates.^11^ Participant-time was only included during continuous registration at the same medical practice to reduce misclassification of event dates.^18^ Missing data were minimal and as such complete-case analysis was performed apart from minor modifications to two confounding variables (substituting practice-level IMD for missing patient-level IMD and defining smoking exposure as the dichotomous ‘never smoked’ or ‘ever smoked’ rather than trying to define ex-smoker status).

### Bias

To reduce bias from health-seeking behaviour, analyses were restricted to individuals previously vaccinated with PPV23 and adjusted for healthcare utilization measures such influenza vaccination uptake and consultation rate at medical practice. Live shingles vaccination was modelled as a time-varying exposure, such that individuals contributed unvaccinated person-time prior to immunisation and vaccinated person-time thereafter to improve comparability between exposed and unexposed person-time and avoid immortal time bias.

### Study size

All eligible participants for the defined study period were included in the analysis.

### Statistical analysis

#### Survival analysis with time-varying vaccine exposure

The study design is summarised in Supplementary Figure 2. Survival analysis with time-varying live shingles vaccine exposure was performed to compare outcome rates during unvaccinated and vaccinated person-time. Individuals entered follow-up at age 69.5 years or on 1^st^ September 2013 (whichever was later). Individuals who subsequently received live shingles vaccination contributed to unvaccinated person-time from study entry until live shingles vaccination and to vaccinated person-time thereafter. Time-zero for the unvaccinated group was study entry, and time-zero for the vaccinated group was receipt of live shingles vaccination and survival curves were constructed using this split person-time.^19^ A study entry of age 69.5 was used because CPRD only provides year of birth and all participants were given the birthday 1^st^ July for analysis purpose. Censoring occurred at death, if participant deregistered from their medical practice, at last data collection date for medical practice, or study end (whichever was earliest). Event-free survival curves were generated using the Simon-Markuch extension of Kaplan–Meier curves to accommodate time-varying vaccination status.^20^

#### Time-to-first event and time-to-subsequent-event modelling

A time-to-first event Cox proportional hazards frailty model with a random intercept for participant medical practice was used to compare the hazard ratio (HR) between vaccinated and unvaccinated person-time for all three outcomes with participants censored when they experienced the outcome event of interest.^21,22^ A second model was used for hospitalisation outcomes which allowed for recurrent events modelling. Time-to-subsequent hospitalisation was modelled using the Andersen-Gill extension of the Cox frailty model in which participants re-entered the study on discharge from hospital and provided an instantaneous risk of subsequent hospitalisation during vaccinated and unvaccinated person-time (Supplementary Figure 2).^23,24^

#### Propensity score overlap weighted pseudo-population

A baseline propensity score was generated for each individual to measure likelihood of that individual going on to receive live shingles vaccination.^17^ The baseline confounding variables described above were inputted into a generalised boosting method (GBM) for propensity score generation in which decision trees fitted over multiple iterations flexibly modelled the relationship between baseline confounding variables and subsequent vaccine exposure.^25,26^ Each individual received a baseline score between zero and one to represent likelihood of subsequent live shingles vaccination. An overlap-weighted pseudo-population was generated using this propensity score by extracting the exposed and unexposed population with overlapping propensity scores (Supplementary Figure 3).^27^ This created an emulated trial scenario in which the risk of exposure in the pseudo-population was balanced between the vaccinated and unvaccinated groups.^27^ All hazards reported in the main results are the vaccine effect in this overlap-weighted pseudo-population. Analyses were conducted in R (version 4.3.0).^28^

#### Pre-defined sensitivity analyses

Exploratory analysis of vaccine effect stratified by sex, age, comorbidity, socioeconomic status, smoking status, and influenza vaccine exposure were performed to assess for potential effect modification and evaluate the need for interaction terms in the model. Vaccine effect with exclusion of population sub-groups was explored including exclusion of frail individuals with death within one year of study entry and exclusion of participants with outcome event associated with varicella-zoster disease event or within 14 days of vaccination. Full description of sensitivity analyses in Supplementary Materials.

## Results

### Baseline characteristics of the overlap-weighted pseudo-population

The CPRD Aurum database included 314,618 participants who had received PPV23 vaccination prior to entry and who were aged 70 years and over between 1^st^ September 2013 and 30^th^ June 2019, giving a total person-time of 1,313,185 years. A total of 174,534 (55%) received the live shingles vaccine during the study period. Participants were registered at 886 medical practices from all ten of England’s 2019 geographical regions. Shingles vaccine coverage ranged from 50% in London to 60% in the East Midlands. The original study population showed differences in age and comorbidity (Supplementary Table 6). As such, an overlap-weighted pseudo-population with balanced risk of exposure was extracted and was used to model risk of outcome associated with live shingles vaccine exposure. Overlap weighting achieved covariate balancing with standardized mean difference <0.1 for all measured covariates and moderate common support with an overlap coefficient of 0.575 (Supplementary Figure 3). The overlap-weighted pseudo-population consisted of 60021 live shingles vaccine unexposed and 60245.6 live shingles vaccine exposed participants with a mean follow-up time of 3.48 (1.96) years and 4.50 (1.53) years respectively. Average time to vaccination from entry in the vaccinated population was 1.88 (1.59) years. Proportion vaccinated in each full study year was similar apart from 2013 and 2019 when only partial year data was available (Supplementary Figure 4). Baseline characteristics of the overlap study population in Table 1.

### Risk of all-cause death, all-cause hospitalisation, and infection-associated hospitalisation associated with live shingles vaccine exposure in the overlap-weighted pseudo-population

The rate of all-cause death in the overlap-weighted pseudo-population was 17.5 per 1000 person-years of observation (pyo) in the live shingles vaccine exposed group compared to 23.3 per 1000 pyo in the vaccine unexposed group. The hazard ratio (HR) of all-cause death following shingle vaccine exposure was 0.64 (95% confidence interval (CI) 0.62, 0.66) (Table 2). The survival curves demonstrate a divergence of survival between one and two years (Figure 1). Proportional hazards assumptions were not violated (Supplementary Figure 5).

The rate of first all-cause hospitalisation in the overlap-weighted pseudo-population was 228 per 1000 pyo in the live shingles vaccine exposed compared to 218 per 1000 pyo in the unexposed with no difference in rate of first hospitalisation detected between groups (HR 0.99 (95% CI 0.97, 1.00)) (Table 2, Figure 1). The Andersen-Gill extension of the Cox model demonstrated a lower instantaneous risk of a subsequent all-cause hospitalisation in individuals with live shingles vaccine exposure (HR 0.84 (95% CI 0.81, 0.88) (Table 3). The rate of first infection-associated hospitalisation was 47.6 per 1000 pyo in live shingles vaccine exposed compared to 52.3 per 1000 pyro in live shingles vaccine unexposed with a lower instantaneous hazard rate of first infection-associated hospitalisation (HR 0.81 (95% CI 0.79, 0.83)) and a lower instantaneous hazard rate of a subsequent infection-associated hospitalisation (HR 0.75 (95% CI 0.73,0.77)). Similar hazard estimates were generated with covariate adjustment and inverse probability weighting (Supplementary Tables 7-10).

### Sensitivity analyses

Sensitivity analyses were modelled using Cox proportional hazards frailty model on the overlap-weighted pseudo-population. No difference in HRs were observed with exclusion of individuals with less than one year follow up; inclusion of individuals with PPV23 after study entry; exclusion of individuals with an outcome within 14 days of live shingles vaccination; nor exclusion of individuals with an outcome within 30 days of a varicella-zoster disease event (Supplementary Tables 11-14). Removal of the older catch-up population, who were offered live shingles vaccination between age 71 and 79 when the vaccine was first rolled out, did not alter the direction of the HR but increased the magnitude of the associated protective effect (Supplementary Table 15). Stratified analyses demonstrated a stronger protective effect associated with vaccine exposure in participants younger at baseline, the most comorbid, the least deprived, and in females (Supplementary Tables 16-18). HR for 70- to 74-year-olds against death was 0.56 (95% CI 0.54, 0.58) versus 0.72 (95% CI 0.68, 0.76) for age 75 years and over, while infection-associated hospitalisation HR for 70- to 74-year-olds was 0.76 (95% CI 0.74, 0.78) versus 0.88 (95% CI 0.85, 0.92) for age 75 years and over. A stronger protective effect was demonstrated in the most comorbid participants against all-cause hospitalisation (HR in least comorbid 1.00 (95% CI 0.98, 1.02) versus most comorbid 0.72 (95% CI 0.57, 0.91)), and infection-associated hospitalisation (HR in least comorbid 0.80 (95% CI 0.78, 0.83) versus most comorbid 0.69 (95% CI 0.52, 0.91)). The least deprived observed a HR of 0.62 (95% CI 0.58, 0.66) against death and 0.79 (95% CI 0.75, 0.83) against infection-associated hospitalisation, compared to 0.72 (95% CI 0.67, 0.78) and 0.88 (95% CI 0.84, 0.93) in the most deprived. Finally, female sex associated with greater vaccine protection (HR for death 0.59 (95% CI 0.56, 0.62) compared to 0.67 (95% CI 0.65, 0.70) for males, and HR for infection-associated hospitalisation 0.78 (95% CI 0.75, 0.80) compared to 0.84 (95% CI 0.81, 0.86) in males).

### Hazard ratio over time since vaccination (post-hoc analysis)

Visualisation of hazard over follow-up time using the time-varying coefficient model confirms the vaccine protective effect is highest within one year of immunisation and attenuates towards the null over follow-up time but persists for at least five years (Supplementary Figure 6).

## Discussion

In this study live shingles vaccine exposure in older adults in England was associated with a 36% reduction in risk of death and a 19% reduction in risk of first infection-associated hospitalisation, which persisted for at least five years following immunisation. Instantaneous risk of a subsequent hospitalisation after discharge was 17% lower in the vaccinated for all-cause hospitalisations, and 25% lower for infection-associated hospitalisation. No protective vaccine effect was detected against first all-cause hospitalisation likely reflecting the inability to distinguish elective from emergency admissions. As elective admissions substantially outnumber emergency admissions in England, any vaccine effect on acute hospitalisations may have been diluted.^29^

As an observational study there is major potential for confounding. Inherent differences in health-seeking behaviours and risk of disease has been described between vaccinated and unvaccinated populations, which can influence observed vaccine effectiveness.^13^ A number of strategies were adopted in this study to account for this including the use of a study population which had all engaged in the UK immunisation schedule by receiving PPV23 immunisation. We created an overlap-weighted pseudo-population aimed at balancing measured baseline characteristics between those who go on to receive vaccination and those who do not. The propensity score used to create the overlap pseudo-population was generated using measures of health status but also measures health-seeking behaviour such as influenza vaccine uptake. The average treatment effect did not differ substantially in direction or magnitude in the overlap-weighted pseudo-population compared to the entire population with covariate adjustment or inverse probability weighting, possibly due to the selection of a population who had already engaged in the UK immunisation schedule.

Live vaccines are not recommended for immunocompromised individuals due to the risk of adverse effects from the vaccine.^14^ While individuals with a defined and recorded immunocompromising condition were excluded from this study, this may not cover individuals who were deemed too vulnerable or frail by the clinician for live immunisation but who do not have a specific immunocompromising condition. Thus, it is possible that the unvaccinated group in this study included more frail individuals. Sensitivity analysis removed individuals who died in the first year of follow-up and were possibly deemed too frail for live shingles immunization which did not alter the estimate substantially. Imbalances at baseline were noted for comorbidity even with overlap weighting. Stratified analysis did not fundamentally alter the associations but indicated that further studies should investigate variability in vaccine effects in relation to social determinants of health and demography. A consistent vaccine effect against all three outcomes was observed across three models (Cox frailty model, Cox-Andersen-Gill frailty model, and time-varying coefficient model).^21^

The study relied on secondary data recorded by healthcare workers for healthcare provision and was not originally intended for use in research. CPRD includes internal checks to ensure validity of data including valid age and date of clinical events.^11^ Date of death recorded in CPRD has been found to largely correspond to Office of National Statistics death records and the broad and non-specific nature of the all-cause hospitalisation outcome limits the risk of misclassification of outcome.^30^ Study follow-up time was limited to when participants were continuously registered at the same GP to limit misclassification of variables. Individuals cannot be linked across practices and although historical clinical information is typically retained following practice registration, some incompleteness may occur. The use of the secondary data source has significant strengths as it facilitates a large sample size from a dataset which is representative of the English population.^11^

This is the first study to evaluate the effect of the live shingles vaccine (*Zostavax®*) in older adults on mortality and hospitalisation. The results are consistent with two recent studies estimating a substantial reduction in dementia diagnosis associated with live shingles vaccination in a Welsh population and a 23% reduction in cardiovascular disease in South Korea which persisted for eight years following immunisation.^8,9,31^ In the case of the dementia association this has also been seen with the recombinant non-live vaccine which may suggest an underlying mechanism linked to protection from the direct consequences of varicella zoster virus, or heterologous immune effects of non-live vaccines.^9,31^ The reduction in cardiovascular deaths was hypothesized as a direct effect through the prevention of varicella-zoster associated cardio-vascular toxicity, however the large effect size raises the possibility of a broader non-specific beneficial effect.

The reduction in infection-associated acute hospitalisations would be consistent with the prevailing ideas on the mechanism of heterologous immune effects, namely programming of the innate and adaptive immune response to provide a more protective and regulated response to inflammation.^32,33^ Whilst there has been extensive study of the varicella zoster targeting immune response of these vaccines, immunological study of off target responses may provide an efficient means of determining whether biologically supportive differences exist post live shingles vaccine.^34^

The findings in this study suggests that despite the impairment of the immune system associated with older age, live vaccines may still elicit heterologous immune-boosting effects which protect from common infectious causes of morbidity and mortality. We cannot exclude residual confounding in this study and therefore doubt will remain about the effect and scale. However, the size of the findings and the emergence of other evidence argues strongly for the need to understand any potential effects better. This study advocates for the incorporation of robust but practical experimental design into vaccine policy and vaccine implementation. For example, phased or cluster randomised roll-out of new vaccines or during age de-escalation to allow robust evidence generation. An immediate question is whether there is any difference between live and recombinant shingles vaccine. The introduction of the two-dose inactivated recombinant Shingrix® vaccine into the UK schedule from 2023 provides a low-cost natural experiment to answer this question and will be immediately relevant for policy.

In conclusion, this study offers evidence for non-specific effects of live shingles vaccine exposure in individuals aged 70-years and over. These findings should be considered when evaluating the cost-effectiveness of live vaccines and shingles vaccination policy in the growing older adult population, and advocates for experimental design in vaccination policy and implementation.

## Data Availability

The datasets used in this study were extracted from CPRD following CPRD approval of the study protocol and through a data sharing agreement between University of Liverpool and CPRD. The same dataset is available from CPRD. The authors are not authorised to share the datasets and are obliged to destroy the datasets according to the data sharing agreement between University of Liverpool and CPRD.

## List of Abbreviations

95%CI: 95% confidence interval
aHR: Adjusted hazard ratio
BCG: Bacillus Calmette–Guérin vaccine
CPRD: Clinical Practice Research Datalink
HES: Health Episode Statistics
HR: Hazard ratio
IMD: Index of Multiple Deprivation
ICD-10: International Statistical Classification of Diseases and Related Health Problems Revision 10
IQR: Interquartile range
NIHR: National Institute of Health Research
PPV23: 23-valent pneumococcal polysaccharide vaccine (Pneumovax™, Merck & Co®)
SD: standard deviation

## Footnotes

## Acknowledgements

This study is based on data from the Clinical Practice Research Datalink (CPRD). CPRD is jointly sponsored by the Medicines and Healthcare products Regulatory Agency and the National Institute for Health Research (NIHR), as part of the Department of Health and Social Care. This work uses data provided by patients and collected by the National Health Service as part of their care and support. We would like to acknowledge all the data providers and general practices that made the anonymised data available for research. This study would not have been possible without the wider CPRD team at University of Liverpool including Pieta Schofield. We are indebted to Professor Peter Smith, London School of Tropical Medicine and Hygiene for making suggestions on earlier drafts of this manuscript. The views expressed are those of the author and not necessarily those of the NIHR, the Department of Health and Social Care or Public Health England.

## Contributors

NF, DH, NB, KD conceived of and designed the study. DH, KD acquired the data; KD analysed the data with support from DH, CJ, LB, MH, NF, and all authors interpreted the output; KD wrote the first draft of the report; and all authors reviewed the draft and final manuscript.

## Consent for publication

Not applicable

## Ethics

The protocol for this study was submitted and approved by the CPRD Independent Scientific Advisory Committee (ISAC). CPRD obtains annual rolling ethical approval (reference number 05/MRE04/87) and no additional ethical approval was required for this project which meets ISAC requirements. The data was accessed via the University of Liverpool CPRD license agreement.

## Availability of data and materials

Data from this study is not available from the authors. Study data is available through formal request from CPRD.

## References

1. Walker JL, Andrews NJ, Amirthalingam G, Forbes H, Langan SM, Thomas SL. Effectiveness of herpes zoster vaccination in an older United Kingdom population. Vaccine. 2018 Apr;36(17):2371–7. doi:10.1016/j.vaccine.2018.02.021

2. Aaby P, Martins CL, Garly ML, Bale C, Andersen A, Rodrigues A, et al. Non-specific effects of standard measles vaccine at 4.5 and 9 months of age on childhood mortality: randomised controlled trial. BMJ. 2010 Nov 30;341(nov30 2):c6495–c6495. doi:10.1136/bmj.c6495

3. Aaby P, Roth A, Ravn H, Napirna BM, Rodrigues A, Lisse IM, et al. Randomized Trial of BCG Vaccination at Birth to Low-Birth-Weight Children: Beneficial Nonspecific Effects in the Neonatal Period? The Journal of Infectious Diseases. 2011 Jul 15;204(2):245–52. doi:10.1093/infdis/jir240

4. Aaby P, Rodrigues A, Biai S, Martins C, Veirum JE, Benn CS, et al. Oral polio vaccination and low case fatality at the paediatric ward in Bissau, Guinea-Bissau. Vaccine. 2004 Aug 13;22(23–24):3014–7. doi:10.1016/j.vaccine.2004.02.009 PubMed PMID: 15297050.

5. de Castro MJ, Pardo-Seco J, Martinón-Torres F. Nonspecific (Heterologous) Protection of Neonatal BCG Vaccination Against Hospitalization Due to Respiratory Infection and Sepsis. Clin Infect Dis. 2015 Jun 1;60(11):1611–9. doi:10.1093/cid/civ144 PubMed PMID: 25725054.

6. Sørup S, Benn CS, Poulsen A, Krause TG, Aaby P, Ravn H. Live vaccine against measles, mumps, and rubella and the risk of hospital admissions for nontargeted infections. JAMA. 2014 Feb 26;311(8):826–35. doi:10.1001/jama.2014.470 PubMed PMID: 24570246.

7. Goodridge HS, Ahmed SS, Curtis N, Kollmann TR, Levy O, Netea MG, et al. Harnessing the beneficial heterologous effects of vaccination. Nature Reviews Immunology. 2016 Jun;16(6):392–400. doi:10.1038/nri.2016.43

8. Lee S, Lee K, Oh J, Kim HJ, Son Y, Kim S, et al. Live zoster vaccination and cardiovascular outcomes: a nationwide, South Korean study. European Heart Journal. 2025 May 5;ehaf230. doi:10.1093/eurheartj/ehaf230

9. Eyting M, Xie M, Michalik F, Heß S, Chung S, Geldsetzer P. A natural experiment on the effect of herpes zoster vaccination on dementia. Nature. 2025 Apr 2;1–9. doi:10.1038/s41586-025-08800-x

10. Stoye G, Warner M, Zaranko B. The past and future of UK health spending [Internet]. Institute for Fiscal Studies; 2024 [cited 2025 Sep 9]. Available from: https://ifs.org.uk/publications/past-and-future-uk-health-spending doi:10.1920/re.ifs.2024.0312

11. Wolf A, Dedman D, Campbell J, Booth H, Lunn D, Chapman J, et al. Data resource profile: Clinical Practice Research Datalink (CPRD) Aurum. Int J Epidemiol. 2019 Dec;48(6):1740–1740g. doi:10.1093/ije/dyz034 PubMed PMID: 30859197; PubMed Central PMCID: PMC6929522.

12. von Elm E, Altman DG, Egger M, Pocock SJ, Gøtzsche PC, Vandenbroucke JP. Strengthening the reporting of observational studies in epidemiology (STROBE) statement: guidelines for reporting observational studies. BMJ. 2007 Oct 20;335(7624):806–8. doi:10.1136/bmj.39335.541782.AD PubMed PMID: 17947786; PubMed Central PMCID: PMC2034723.

13. Weinberg GA, Szilagyi PG. Vaccine Epidemiology: Efficacy, Effectiveness, and the Translational Research Roadmap. The Journal of Infectious Diseases. 2010 Jun 1;201(11):1607–10. doi:10.1086/652404

14. UKHSA UHSA. GOV.UK [Internet]. 2021 [cited 2025 Aug 12]. Immunisation against infectious disease. The Green Book. Available from: https://www.gov.uk/government/collections/immunisation-against-infectious-disease-the-green-book

15. Torisson G, Rosenqvist M, Melander O, Resman F. Hospitalisations with infectious disease diagnoses in somatic healthcare between 1998 and 2019: A nationwide, register-based study in Swedish adults. The Lancet Regional Health – Europe. 2022 May 1;16. doi:10.1016/j.lanepe.2022.100343 PubMed PMID: 35360441.

16. Quan H, Li B, Couris CM, Fushimi K, Graham P, Hider P, et al. Updating and validating the Charlson comorbidity index and score for risk adjustment in hospital discharge abstracts using data from 6 countries. Am J Epidemiol. 2011 Mar 15;173(6):676–82. doi:10.1093/aje/kwq433 PubMed PMID: 21330339.

17. Austin PC. An Introduction to Propensity Score Methods for Reducing the Effects of Confounding in Observational Studies. Multivariate Behavioral Research. 2011 May 31;46(3):399–424. doi:10.1080/00273171.2011.568786 PubMed PMID: 21818162.

18. Lewis JD, Bilker WB, Weinstein RB, Strom BL. The relationship between time since registration and measured incidence rates in the General Practice Research Database. Pharmacoepidemiol Drug Saf. 2005 Jul;14(7):443–51. doi:10.1002/pds.1115 PubMed PMID: 15898131.

19. Zhang Z, Reinikainen J, Adeleke KA, Pieterse ME, Groothuis-Oudshoorn CGM. Time-varying covariates and coefficients in Cox regression models. Ann Transl Med. 2018 Apr;6(7):121. doi:10.21037/atm.2018.02.12 PubMed PMID: 29955581; PubMed Central PMCID: PMC6015946.

20. Schultz LR, Peterson EL, Breslau N. Graphing survival curve estimates for time-dependent covariates. Int J Methods Psychiatr Res. 2002;11(2):68–74. doi:10.1002/mpr.124 PubMed PMID: 12459796; PubMed Central PMCID: PMC6878542.

21. Therneau T, Crowson C, Atkinson E. ResearchGate [Internet]. 2024 [cited 2025 Jul 24]. Using Time Dependent Covariates and Time Dependent Coefficients in the Cox Model. Available from: https://www.researchgate.net/publication/265041278_Using_Time_Dependent_Covariates_and_Time_Dependent_Coefficients_in_the_Cox_Model

22. Therneau TM, Grambsch PM, Pankratz VS. Penalized Survival Models and Frailty. Journal of Computational and Graphical Statistics. 2003;12(1):156–75.

23. Andersen P, Gill R. Cox’s Regression Model for Counting Processes: A Large Sample Study. The Annals of Statistics. 1982 Dec 1;10. doi:10.1214/aos/1176345976

24. Bonnett LJ, Spain T, Hunt A, Hutton JL, Watson V, Marson AG, et al. Guide to evaluating performance of prediction models for recurrent clinical events. Diagn Progn Res. 2025 Mar 17;9:6. doi:10.1186/s41512-025-00187-7 PubMed PMID: 40098007; PubMed Central PMCID: PMC11912649.

25. Garrido MM, Kelley AS, Paris J, Roza K, Meier DE, Morrison RS, et al. Methods for Constructing and Assessing Propensity Scores. Health Services Research. 2014;49(5):1701–20. doi: 10.1111/1475-6773.12182

26. Olmos A, Govindasamy P. Propensity Scores: A Practical Introduction Using R. Journal of MultiDisciplinary Evaluation. 2015 Oct 12;11(25):25.

27. Cao Z, Ghazi L, Mastrogiacomo C, Forastiere L, Wilson FP, Li F. Using overlap weights to address extreme propensity scores in estimating restricted mean counterfactual survival times. Am J Epidemiol. 2025 Aug 5;194(8):2402–11. doi:10.1093/aje/kwae416 PubMed PMID: 39489504; PubMed Central PMCID: PMC12342919.

28. R Core Team. R: The R Project for Statistical Computing [Internet]. [cited 2025 Oct 23]. Available from: https://www.r-project.org/

29. NHS England Digital [Internet]. [cited 2026 Apr 9]. Summary Report - Admissions. Available from: https://digital.nhs.uk/data-and-information/publications/statistical/hospital-admitted-patient-care-activity/2019-20/summary-reports---apc---admissions

30. Gallagher AM, Dedman D, Padmanabhan S, Leufkens HGM, Vries F de. The accuracy of date of death recording in the Clinical Practice Research Datalink GOLD database in England compared with the Office for National Statistics death registrations. Pharmacoepidemiology and Drug Safety. 2019;28(5):563–9. doi:10.1002/pds.4747

31. Taquet M, Dercon Q, Todd JA, Harrison PJ. The recombinant shingles vaccine is associated with lower risk of dementia. Nat Med. 2024 Oct;30(10):2777–81. doi:10.1038/s41591-024-03201-5

32. Kleinnijenhuis J, Quintin J, Preijers F, Benn CS, Joosten LAB, Jacobs C, et al. Long-Lasting Effects of BCG Vaccination on Both Heterologous Th1/Th17 Responses and Innate Trained Immunity. JIN. 2014;6(2):152–8. doi:10.1159/000355628 PubMed PMID: 24192057.

33. Kleinnijenhuis J, Quintin J, Preijers F, Joosten LAB, Jacobs C, Xavier RJ, et al. BCG-induced trained immunity in NK cells: role for non-specific protection to infection. Clin Immunol. 2014 Dec;155(2):213–9. doi:10.1016/j.clim.2014.10.005 PubMed PMID: 25451159; PubMed Central PMCID: PMC5084088.

34. Sei JJ, Cox KS, Dubey SA, Antonello JM, Krah DL, Casimiro DR, et al. Effector and Central Memory Poly-Functional CD4+ and CD8+ T Cells are Boosted upon ZOSTAVAX® Vaccination. Frontiers in Immunology [Internet]. 2015 [cited 2023 Sep 15];6. Available from: https://www.frontiersin.org/articles/10.3389/fimmu.2015.00553

