## Supplementary materials for "Evidence for non-specific effects of live Shingles vaccination against all-cause death and hospitalisation in older adults in England: a population-based cohort study"

#### **Supplementary table 1: STROBE checklist**

| **Item No.** | **Recommendation** | **Location in Manuscript** |
| --- | --- | --- |
| **TITLE & ABSTRACT** | | |
| 1 | Indicate the study’s design in the title or abstract | Title; Abstract |
| **INTRODUCTION** | | |
| 2 | Explain the scientific background and rationale | Background |
| 3 | State specific objectives, including any prespecified hypotheses | End of Background |
| **METHODS** |  |  |
| 4 | Present key elements of study design early | Methods – Study design and data source |
| 5 | Describe the setting, locations, and relevant dates, including recruitment, exposure, follow-up, and data collection periods | Methods – Study setting |
| 6a | Give the eligibility criteria, and sources and methods of participant selection | Methods – Study participants  Supplementary Methodology – Study population |
| 6b | Describe methods of follow-up | Methods – Statistical analysis |
| 7 | Clearly define all outcomes, exposures, predictors, potential confounders, and effect modifiers | Methods – Primary exposure variable; Outcome variables; Confounding variables |
| 8 | For each variable of interest, give sources of data and measurement methods | Methods – Primary exposure variable; Confounding variables; Outcome variables; Data collection and measurement  Supplementary Methodology – Propensity Score Generation |
| 9 | Describe any efforts to address potential sources of bias | Methods – Study participants; Bias |
| 10 | Explain how the study size was arrived at | Methods – Study size |
| 11 | Explain how quantitative variables were handled (including any groupings) | Methods – Confounding variables; Statistical analysis |
| 12a | Describe all statistical methods, including those used to control for confounding | Methods – Statistical analysis |
| 12b | Describe methods for examining subgroups and interactions | Methods – Statistical analysis |
| 12c | Explain how missing data were addressed | Methods – Data collection and measurement |
| 12d | If applicable, explain how loss to follow-up was addressed | Methods – Statistical analysis |
| 12e | Describe any sensitivity analyses | Methods – Statistical analysis  Supplementary Methodology – Supplementary Analyses |
| **RESULTS** |  |  |
| 13a | Report numbers of individuals at each stage (e.g., eligibility, included, analysed) | Results – Study population; Supplementary Figure 1 |
| 13b | Give reasons for non-participation at each stage | Supplementary Figure 1 |
| 13c | Consider use of a flow diagram | Supplementary Figure 1 |
| 14a | Give characteristics of study participants (demographic, clinical, social) | Results – Study population; Table 1 |
| 14b | Indicate number of participants with missing data for each variable of interest | Table 1 |
| 14c | Summarise follow-up time | Results |
| 15 | Report numbers of outcome events or summary measures | Table 2; Table 3; Figure 1 |
| 16a | Provide unadjusted and adjusted estimates with precision (e.g., 95% CI); make clear which confounders were adjusted for | Results; Table 2, Table 3 |
| 16b | Report category boundaries when continuous variables were categorised | Methods – Confounding variables  Supplementary Methodology: Propensity Score Generation |
| 16c | If relevant, consider translating relative risks into absolute risks | Results Supplementary Results – Absolute measures of survival |
| 17 | Report other analyses (e.g., subgroup, sensitivity, interactions) | Results  Supplementary Results |
| **DISCUSSION** |  |  |
| 18 | Summarise key results with reference to objectives | Discussion – Paragraph 1 |
| 19 | Discuss limitations, including potential for bias or imprecision | Discussion – Paragraph 2, 3, 4 |
| 20 | Discuss generalisability | Discussion – Paragraph 5, 6, 7 |
| **OTHER INFORMATION** |  |  |
| 21 | Provide source of funding and role of funders (if any) | Footnotes |
| 22 | Ethical approval statement | Footnotes |

### **1. SUPPLEMENTARY METHODOLOGY: STUDY POPULATION**

#### **Study population exclusion criteria**

Participants were excluded if they had received the lower-dose varicella zoster vaccine (*Varivax®*) (Supplementary Table 5); if they have received multiple doses of the live shingles vaccine which might reflect data inconsistencies or an unusual patient cohort; if they had received the live shingles vaccine prior to age 70 years; or if they had received the live shingles vaccine prior to September 2013. Participants were excluded if they received Zostavax before the start of eligible follow-up. Start of eligible follow-up was defined as 28 weeks after the date of continuous registration at their medical practice to reduce inclusion of historical vaccination records recorded after registration (i.e., vaccination records entered retrospectively at the time of registration rather than reflecting new vaccination events).^1^ Participants with inconsistent or implausible events, for example events occurring after death, were excluded but this occurred infrequently (Supplementary Figure 1).

#### **Supplementary table 2: Medcodes to define immunosuppression**

| medcode | readterm |
| --- | --- |
| 47106 | Patient immunocompromised |
| 22307 | Patient immunosuppressed |
| 34310 | Total excision of spleen |
| 108246 | Total excision of spleen and replantation of fragments |
| 27514 | Total splenectomy |
| 1393 | Splenectomy NEC |
| 31041 | Laparoscopic total splenectomy |
| 102966 | Other specified total excision of spleen |
| 59784 | Total excision of spleen NOS |
| 106015 | Transplantation of spleen |
| 32133 | Botulism |
| 65460 | Malignant neoplasm of spleen NEC |
| 108667 | Angiosarcoma of spleen |
| 72224 | Fibrosarcoma of spleen |
| 93778 | Malignant neoplasm of spleen NOS |
| 12323 | Malignant neoplasm of lymphatic and haemopoietic tissue |
| 41369 | Lymphosarcoma and reticulosarcoma |
| 1481 | Reticulosarcoma |
| 60242 | Reticulosarcoma of unspecified site |
| 71031 | Reticulosarcoma of lymph nodes of head, face and neck |
| 70374 | Reticulosarcoma of intra-abdominal lymph nodes |
| 95058 | Reticulosarcoma of spleen |
| 99240 | Reticulosarcoma NOS |
| 27416 | Lymphosarcoma |
| 71625 | Lymphosarcoma of unspecified site |
| 71238 | Lymphosarcoma of lymph nodes of head, face and neck |
| 62380 | Lymphosarcoma of intrathoracic lymph nodes |
| 64670 | Lymphosarcoma of intra-abdominal lymph nodes |
| 100352 | Lymphosarcoma of lymph nodes of inguinal region and leg |
| 103245 | Lymphosarcoma of spleen |
| 104790 | Lymphosarcoma of lymph nodes of multiple sites |
| 63723 | Lymphosarcoma NOS |
| 21402 | Burkitt's lymphoma |
| 59115 | Burkitt's lymphoma of lymph nodes of head, face and neck |
| 100006 | Burkitt's lymphoma of intrathoracic lymph nodes |
| 97577 | Burkitt's lymphoma of intra-abdominal lymph nodes |
| 92380 | Burkitt's lymphoma of lymph nodes of inguinal region and leg |
| 71304 | Burkitt's lymphoma NOS |
| 99887 | Other specified reticulosarcoma or lymphosarcoma |
| 99951 | Reticulosarcoma or lymphosarcoma NOS |
| 2462 | Hodgkin's disease |
| 65489 | Hodgkin's paragranuloma |
| 100423 | Hodgkin's paragranuloma of lymph nodes of head, face, neck |
| 98840 | Hodgkin's paragranuloma of intra-abdominal lymph nodes |
| 44196 | Hodgkin's granuloma |
| 98909 | Hodgkin's granuloma of lymph nodes of head, face and neck |
| 64036 | Hodgkin's sarcoma |
| 68039 | Hodgkin's sarcoma of lymph nodes of axilla and upper limb |
| 38939 | Hodgkin's disease, lymphocytic-histiocytic predominance |
| 71142 | Hodgkin's, lymphocytic-histiocytic predominance unspec site |
| 68330 | Hodgkin's, lymphocytic-histiocytic pred of head, face, neck |
| 92245 | Hodgkin's, lymphocytic-histiocytic pred intrathoracic nodes |
| 73532 | Hodgkin's, lymphocytic-histiocytic pred intra-abdominal node |
| 93951 | Hodgkin's, lymphocytic-histiocytic pred inguinal and leg |
| 95338 | Hodgkin's, lymphocytic-histiocytic pred intrapelvic nodes |
| 106911 | Hodgkin's, lymphocytic-histiocytic predominance of spleen |
| 104743 | Hodgkin's, lymphocytic-histiocytic pred of multiple sites |
| 29876 | Hodgkin's, lymphocytic-histiocytic predominance NOS |
| 29178 | Hodgkin's disease, nodular sclerosis |
| 57225 | Hodgkin's disease, nodular sclerosis of unspecified site |
| 55303 | Hodgkin's nodular sclerosis of head, face and neck |
| 67506 | Hodgkin's nodular sclerosis of intrathoracic lymph nodes |
| 61149 | Hodgkin's nodular sclerosis of intra-abdominal lymph nodes |
| 65483 | Hodgkin's nodular sclerosis of lymph nodes of axilla and arm |
| 105472 | Hodgkin's disease, nodular sclerosis of spleen |
| 19140 | Hodgkin's nodular sclerosis of lymph nodes of multiple sites |
| 63054 | Hodgkin's disease, nodular sclerosis NOS |
| 49605 | Hodgkin's disease, mixed cellularity |
| 97863 | Hodgkin's disease, mixed cellularity of unspecified site |
| 94407 | Hodgkin's mixed cellularity of lymph nodes head, face, neck |
| 58684 | Hodgkin's mixed cellularity of intrathoracic lymph nodes |
| 108886 | Hodgkin's mixed cellularity of lymph nodes inguinal and leg |
| 94005 | Hodgkin's disease, mixed cellularity NOS |
| 67703 | Hodgkin's disease, lymphocytic depletion |
| 95049 | Hodgkin's lymphocytic depletion of unspecified site |
| 111942 | Hodgkin's lymphocytic depletion of head, face and neck |
| 63625 | Hodgkin's lymphocytic depletion lymph nodes axilla and arm |
| 110563 | Hodgkin's lymphocytic depletion lymph nodes inguinal and leg |
| 101715 | Hodgkin's disease, lymphocytic depletion of spleen |
| 107032 | Hodgkin's lymphocytic depletion lymph nodes multiple sites |
| 101530 | Hodgkin's disease, lymphocytic depletion NOS |
| 104895 | Nodular lymphocyte predominant Hodgkin lymphoma |
| 105841 | Nodular sclerosis classical Hodgkin lymphoma |
| 108775 | Mixed cellularity classical Hodgkin lymphoma |
| 106597 | Lymphocyte-rich classical Hodgkin lymphoma |
| 104484 | Other classical Hodgkin lymphoma |
| 53397 | Hodgkin's disease NOS |
| 61662 | Hodgkin's disease NOS, unspecified site |
| 59778 | Hodgkin's disease NOS of lymph nodes of head, face and neck |
| 59755 | Hodgkin's disease NOS of intrathoracic lymph nodes |
| 107804 | Hodgkin's disease NOS of intra-abdominal lymph nodes |
| 91900 | Hodgkin's disease NOS of lymph nodes of axilla and arm |
| 99012 | Hodgkin's disease NOS of lymph nodes inguinal region and leg |
| 94279 | Hodgkin's disease NOS of spleen |
| 97746 | Hodgkin's disease NOS of lymph nodes of multiple sites |
| 42461 | Hodgkin's disease NOS |
| 5179 | Nodular lymphoma (Brill - Symmers disease) |
| 66327 | Nodular lymphoma of unspecified site |
| 45264 | Nodular lymphoma of lymph nodes of head, face and neck |
| 105203 | Nodular lymphoma of intrathoracic lymph nodes |
| 92068 | Nodular lymphoma of intra-abdominal lymph nodes |
| 111766 | Nodular lymphoma of lymph nodes of axilla and upper limb |
| 94995 | Nodular lymphoma of lymph nodes of inguinal region and leg |
| 58082 | Nodular lymphoma of lymph nodes of multiple sites |
| 65701 | Nodular lymphoma NOS |
| 12006 | Mycosis fungoides |
| 95949 | Mycosis fungoides of unspecified site |
| 91674 | Mycosis fungoides of intra-abdominal lymph nodes |
| 96379 | Mycosis fungoides of lymph nodes of axilla and upper limb |
| 72714 | Mycosis fungoides of lymph nodes of inguinal region and leg |
| 95012 | Mycosis fungoides of lymph nodes of multiple sites |
| 38005 | Mycosis fungoides NOS |
| 35014 | Sezary's disease |
| 100532 | Sezary's disease NOS |
| 27330 | Leukaemic reticuloendotheliosis |
| 65122 | Leukaemic reticuloendotheliosis of unspecified sites |
| 65123 | Leukaemic reticuloend of intra-abdominal lymph nodes |
| 73777 | Leukaemic reticuloendotheliosis NOS |
| 34926 | Letterer-Siwe disease |
| 102715 | Letterer-Siwe disease of unspecified sites |
| 102158 | Letterer-Siwe disease of intrathoracic lymph nodes |
| 54083 | Letterer-Siwe disease of lymph nodes of multiple sites |
| 47204 | Letterer-Siwe disease NOS |
| 15036 | Malignant mast cell tumours |
| 103900 | Mast cell malignancy of unspecified site |
| 100615 | Mast cell malignancy of lymph nodes inguinal region and leg |
| 31324 | Mast cell malignancy of lymph nodes of multiple sites |
| 89657 | Malignant mast cell tumour NOS |
| 3604 | Non - Hodgkin's lymphoma |
| 28639 | Follicular non-Hodgkin's small cleaved cell lymphoma |
| 70842 | Follicular non-Hodg mixed sml cleavd & lge cell lymphoma |
| 49262 | Follicular non-Hodgkin's large cell lymphoma |
| 50668 | Diffuse non-Hodgkin's small cell (diffuse) lymphoma |
| 108182 | Diffuse non-Hodgkin's small cleaved cell (diffuse) lymphoma |
| 50695 | Diffuse non-Hodgkin mixed sml & lge cell (diffuse) lymphoma |
| 53551 | Diffuse non-Hodgkin's immunoblastic (diffuse) lymphoma |
| 17460 | Diffuse non-Hodgkin's lymphoblastic (diffuse) lymphoma |
| 65180 | Diffuse non-Hodgkin's lymphoma undifferentiated (diffuse) |
| 95715 | Mucosa-associated lymphoma |
| 101114 | Diffuse non-Hodgkin's large cell lymphoma |
| 31576 | Other types of follicular non-Hodgkin's lymphoma |
| 21549 | Follicular non-Hodgkin's lymphoma |
| 70509 | Diffuse non-Hodgkin's centroblastic lymphoma |
| 102594 | Diffuse large B-cell lymphoma |
| 105966 | Extranod marg zone B-cell lymphom mucosa-assoc lymphoid tiss |
| 105038 | Mediastinal (thymic) large B-cell lymphoma |
| 31794 | Unspecified B-cell non-Hodgkin's lymphoma |
| 39798 | Diffuse non-Hodgkin's lymphoma, unspecified |
| 104152 | Follicular lymphoma |
| 105889 | Follicular lymphoma grade 1 |
| 105095 | Follicular lymphoma grade 2 |
| 107166 | Follicular lymphoma grade 3 |
| 105020 | Follicular lymphoma grade 3a |
| 107973 | Follicular lymphoma grade 3b |
| 106969 | Diffuse follicle centre lymphoma |
| 108719 | Cutaneous follicle centre lymphoma |
| 106063 | Other types of follicular lymphoma |
| 105792 | Multifocal multisystemic dissem Langerhans-cell histiocytosi |
| 105335 | Sarcoma of dendritic cells |
| 105559 | Anaplastic large cell lymphoma, ALK-positive |
| 105955 | Anaplastic large cell lymphoma, ALK-negative |
| 109780 | Extranodal NK/T-cell lymphoma, nasal type |
| 107949 | Hepatosplenic T-cell lymphoma |
| 105709 | Enteropathy-associated T-cell lymphoma |
| 105925 | Subcutaneous panniculitic T-cell lymphoma |
| 105375 | Blastic NK-cell lymphoma |
| 105636 | Angioimmunoblastic T-cell lymphoma |
| 105286 | Primary cutaneous CD30-positive T-cell proliferations |
| 104934 | Other mature T/NK-cell lymphoma |
| 106884 | Nonfollicular lymphoma |
| 104386 | Small cell B-cell lymphoma |
| 104620 | Mantle cell lymphoma |
| 104412 | Lymphoblastic (diffuse) lymphoma |
| 111682 | Other non-follicular lymphoma |
| 17887 | Malignant lymphoma otherwise specified |
| 90201 | T-zone lymphoma |
| 57737 | Lymphoepithelioid lymphoma |
| 12464 | Peripheral T-cell lymphoma |
| 44318 | Oth and unspecif peripheral & cutaneous T-cell lymphomas |
| 12335 | Malignant lymphoma NOS |
| 57427 | Malignant lymphoma NOS of unspecified site |
| 50696 | Malignant lymphoma NOS of lymph nodes of head, face and neck |
| 72725 | Malignant lymphoma NOS of intrathoracic lymph nodes |
| 42579 | Malignant lymphoma NOS of intra-abdominal lymph nodes |
| 34089 | Malignant lymphoma NOS of lymph nodes of axilla and arm |
| 63105 | Malignant lymphoma NOS of lymph node inguinal region and leg |
| 71262 | Malignant lymphoma NOS of intrapelvic lymph nodes |
| 60092 | Malignant lymphoma NOS of spleen |
| 15504 | Malignant lymphoma NOS of lymph nodes of multiple sites |
| 15027 | Malignant lymphoma NOS |
| 65434 | Malignant neoplasms of lymphoid and histiocytic tissue NOS |
| 37182 | Multiple myeloma and immunoproliferative neoplasms |
| 4944 | Multiple myeloma |
| 22158 | Malignant plasma cell neoplasm, extramedullary plasmacytoma |
| 19028 | Solitary myeloma |
| 21329 | Plasmacytoma NOS |
| 46042 | Lambda light chain myeloma |
| 104418 | Solitary plasmacytoma |
| 39187 | Plasma cell leukaemia |
| 64567 | Other immunoproliferative neoplasms |
| 43450 | Immunoproliferative neoplasm or myeloma NOS |
| 19372 | Lymphoid leukaemia |
| 4251 | Acute lymphoid leukaemia |
| 104325 | B-cell acute lymphoblastic leukaemia |
| 8625 | Chronic lymphoid leukaemia |
| 104328 | B-cell chronic lymphocytic leukaemia |
| 107052 | Clinical stage A chronic lymphocytic leukaemia |
| 106924 | Clinical stage B chronic lymphocytic leukaemia |
| 107163 | Clinical stage C chronic lymphocytic leukaemia |
| 72774 | Subacute lymphoid leukaemia |
| 49725 | Other lymphoid leukaemia |
| 31586 | Prolymphocytic leukaemia |
| 37461 | Adult T-cell leukaemia |
| 108656 | B-cell prolymphocytic leukaemia |
| 107643 | T-cell prolymphocytic leukaemia |
| 104939 | Adult T-cell lymphoma/leukaemia (HTLV-1-associated) |
| 38331 | Other lymphoid leukaemia NOS |
| 38914 | Lymphoid leukaemia NOS |
| 7176 | Myeloid leukaemia |
| 4413 | Acute myeloid leukaemia |
| 10726 | Chronic myeloid leukaemia |
| 100786 | Chronic eosinophilic leukaemia |
| 105957 | Chronic myeloid leukaemia, BCR/ABL positive |
| 102783 | Chronic neutrophilic leukaemia |
| 107236 | Atypical chronic myeloid leukaemia, BCR/ABL negative |
| 27520 | Chronic myeloid leukaemia NOS |
| 63475 | Subacute myeloid leukaemia |
| 70724 | Myeloid sarcoma |
| 52327 | Chloroma |
| 39629 | Granulocytic sarcoma |
| 104788 | Acute myeloblastic leukaemia |
| 112440 | Other myeloid leukaemia |
| 27664 | Acute promyelocytic leukaemia |
| 66089 | Other myeloid leukaemia NOS |
| 33344 | Myeloid leukaemia NOS |
| 35875 | Monocytic leukaemia |
| 19974 | Acute monocytic leukaemia |
| 27458 | Chronic monocytic leukaemia |
| 101606 | Subacute monocytic leukaemia |
| 108424 | Acute monoblastic leukaemia |
| 99015 | Other monocytic leukaemia |
| 103645 | Other monocytic leukaemia NOS |
| 93342 | Monocytic leukaemia NOS |
| 37272 | Other specified leukaemia |
| 42539 | Acute erythraemia and erythroleukaemia |
| 37468 | Chronic erythraemia |
| 57671 | Megakaryocytic leukaemia |
| 65721 | Mast cell leukaemia |
| 50858 | Acute panmyelosis |
| 28276 | Acute myelofibrosis |
| 110838 | Acute erythroid leukaemia |
| 104273 | Myelodysplastic and myeloproliferative disease |
| 94174 | Other and unspecified leukaemia |
| 72197 | Lymphosarcoma cell leukaemia |
| 99413 | Other and unspecified leukaemia NOS |
| 30632 | Other specified leukaemia NOS |
| 25191 | Leukaemia of unspecified cell type |
| 4072 | Acute leukaemia NOS |
| 16416 | Chronic leukaemia NOS |
| 54793 | Subacute leukaemia NOS |
| 34692 | Other leukaemia of unspecified cell type |
| 4250 | Leukaemia NOS |
| 20440 | Myelomonocytic leukaemia |
| 61500 | Acute myelomonocytic leukaemia |
| 22050 | Chronic myelomonocytic leukaemia |
| 104475 | Subacute myelomonocytic leukaemia |
| 105069 | Juvenile myelomonocytic leukaemia |
| 30646 | Malignant neoplasm lymphatic or haematopoietic tissue OS |
| 6115 | Myeloproliferative disorder |
| 39336 | Myelosclerosis with myeloid metaplasia |
| 49301 | Malignant neoplasm lymphatic or haematopoietic tissue NOS |
| 25493 | Neoplasm of uncertain behaviour of spleen |
| 2481 | Polycythaemia vera |
| 11950 | Essential (haemorrhagic) thrombocythaemia |
| 106993 | Refractory anaemia with ring sideroblasts |
| 104740 | Refractory anaemia with multilineage dysplasia |
| 105915 | 5Q minus syndrome |
| 45285 | Myelodysplastic syndrome, unspecified |
| 46444 | [M]Erythroleukaemias |
| 70935 | [M]Erythroleukaemia |
| 40740 | [X]Malignant neoplasms of lymphoid, haematopoietic and rela |
| 43415 | [X]Other Hodgkin's disease |
| 67518 | [X]Other types of follicular non-Hodgkin's lymphoma |
| 98596 | [X]Other types of diffuse non-Hodgkin's lymphoma |
| 64336 | [X]Other specified types of non-Hodgkin's lymphoma |
| 102688 | [X]Other malignant immunoproliferative diseases |
| 67029 | [X]Other lymphoid leukaemia |
| 61693 | [X]Other myeloid leukaemia |
| 89762 | [X]Other monocytic leukaemia |
| 89329 | [X]Other specified leukaemias |
| 65165 | [X]Other leukaemia of unspecified cell type |
| 105025 | [X]Oth spcf mal neoplsm/lymphoid,haematopoietic+rltd tissue |
| 72500 | [X]Mal neoplasm/lymphoid,haematopoietic+related tissu,unspcf |
| 64515 | [X]Diffuse non-Hodgkin's lymphoma, unspecified |
| 109714 | [X]Oth and unspecif peripheral & cutaneous T-cell lymphomas |
| 63375 | [X]Unspecified B-cell non-Hodgkin's lymphoma |
| 8649 | [X]Non-Hodgkin's lymphoma, unspecified type |
| 45143 | [X]Myelodysplastic syndrome, unspecified |
| 69545 | Chediak-Higashi syndrome |
| 15883 | Monoclonal paraproteinaemia |
| 106381 | Monoclonal gammopathy of uncertain significance |
| 3451 | Other paraproteinaemias |
| 12306 | Benign paraproteinaemia |
| 12386 | Paraproteinaemia NOS |
| 16527 | Macroglobulinaemia |
| 10411 | Waldenstrom's macroglobulinaemia |
| 101350 | Alpha heavy chain disease |
| 99067 | Gamma heavy chain disease |
| 108102 | Heavy chain disease |
| 71994 | Macroglobulinaemia NOS |
| 18486 | Hereditary angio-oedema |
| 51041 | Haemophagocytic lymphohistiocytosis |
| 27934 | Haemophagocytic syndrome, infection-associated |
| 104554 | Tumour lysis syndrome |
| 110183 | Macrophage activation syndrome |
| 44147 | Deficiencies of humoral immunity |
| 15137 | Hypogammaglobulinaemia NOS |
| 8548 | Selective IgA immunodeficiency |
| 18701 | Selective IgM immunodeficiency |
| 18700 | Selective IgG immunodeficiency |
| 68440 | Other selective immunoglobulin deficiency |
| 57161 | Congenital hypogammaglobulinaemia |
| 69373 | Immunodeficiency with IgM hypergammaglobulinaemia |
| 57322 | Common variable immunodeficiency |
| 92569 | Transient infant hypogammaglobulinaemia |
| 62598 | Agammaglobulinaemia NEC |
| 69184 | Dysimmunoglobulinaemia NEC |
| 60026 | Antibod def wth nr-norm imunoglob/or wth hyperimunoglobaemia |
| 69854 | Other specified deficiency of humoral immunity |
| 16295 | Deficiency of humoral immunity NOS |
| 48307 | Deficiencies of cell-mediated immunity |
| 50665 | Predominantly T-cell immuno-deficiency NOS |
| 31322 | Wiskott - Aldrich syndrome |
| 62236 | Combined immunity deficiency |
| 48293 | Severe combined immunodeficiency |
| 31541 | Severe combined immunodefiency with reticular dysgenesis |
| 66073 | Severe combined immunodef with low T- and B-cell numbers |
| 49542 | Severe combined immunodef with low or normal B-cell numbers |
| 50526 | Major histocompatibility complex class I deficiency |
| 103977 | Major histocompatibility complex class II deficiency |
| 62328 | Combined immunity deficiency NOS |
| 3129 | Unspecified immunity deficiency |
| 65617 | Immunodeficiency with short-limbed stature |
| 56108 | Immunodef follow hereditary defect respon Epstein-Barr vir |
| 54203 | Hyperimmunoglobulin E syndrome |
| 21975 | Common variable immunodeficiency |
| 93892 | Com var immunodef with predom abn B-cell numbers and functns |
| 66857 | Common variable immunodef wth autoantibod to B- or T-cells |
| 36408 | Defects in the complement system |
| 109671 | Mannose-binding lectin deficiency |
| 54904 | Lymphocyte function antigen-1 defect |
| 63204 | Idiopathic agranulocytosis |
| 32141 | Cyclical neutropenia |
| 91911 | Functional disorders of polymorphonuclear neutrophils |
| 66049 | Congenital dysphagocytosis |
| 40950 | Familial erythrophagocytic lymph histiocytosis |
| 18781 | Chronic granulomatous disease |
| 38306 | Polymorphonuclear neutrophil disorder NOS |
| 39034 | Hyposplenism |
| 34150 | Hypergammaglobulinaemia |
| 5572 | Myelofibrosis |
| 2337 | Pseudocholinesterase deficiency |
| 73583 | Ataxia-telangiectasia |
| 12699 | Myoneural disorders |
| 5655 | Myasthenia gravis |
| 66740 | Juvenile or adult myasthenia gravis |
| 27515 | Myasthenia gravis NOS |
| 51640 | Myasthenic syndrome due to disease EC |
| 32270 | Eaton-Lambert syndrome |
| 57551 | Myasthenic syndrome due to other malignancy |
| 95005 | Myasthenic syndrome due to botulism |
| 39420 | Myasthenic syndrome due to diabetic amyotrophy |
| 61069 | Myasthenic syndrome due to hypothyroidism |
| 56973 | Myasthenic syndrome due to pernicious anaemia |
| 47695 | Myasthenic syndrome due to thyrotoxicosis |
| 65825 | Myasthenic syndrome due to disease NOS |
| 63323 | Toxic myoneural disorder |
| 53317 | Congenital and developmental myasthenia |
| 20261 | Other specific myoneural disorder |
| 11346 | Myoneural disorder NOS |
| 100043 | [X]Myasthenic syndromes/other diseases classified elsewhere |
| 52604 | Lethal midline granuloma |
| 57525 | Leiner's disease |
| 40406 | Poikiloderma vasculare atrophicans |
| 70236 | Bloom syndrome |
| 62041 | Shwachman-Diamond syndrome |
| 22319 | Absent spleen |
| 101375 | Acquired C1 esterase inhibitor deficiency |
| 101075 | Hereditary C1 esterase inhibitor deficiency |
| 4072 | Acute leukaemia NOS |
| 4251 | Acute lymphoid leukaemia |
| 19974 | Acute monocytic leukaemia |
| 4413 | Acute myeloid leukaemia |
| 50858 | Acute panmyelosis |
| 27664 | Acute promyelocytic leukaemia |
| 37461 | Adult T-cell leukaemia |
| 52327 | Chloroma |
| 100786 | Chronic eosinophilic leukaemia |
| 16416 | Chronic leukaemia NOS |
| 8625 | Chronic lymphoid leukaemia |
| 27458 | Chronic monocytic leukaemia |
| 10726 | Chronic myeloid leukaemia |
| 27520 | Chronic myeloid leukaemia NOS |
| 22050 | Chronic myelomonocytic leukaemia |
| 102783 | Chronic neutrophilic leukaemia |
| 50668 | Diffuse non-Hodgkin's small cell (diffuse) lymphoma |
| 53551 | Diffuse non-Hodgkin's immunoblastic (diffuse) lymphoma |
| 17460 | Diffuse non-Hodgkin's lymphoblastic (diffuse) lymphoma |
| 65180 | Diffuse non-Hodgkin's lymphoma undifferentiated (diffuse) |
| 21549 | Follicular non-Hodgkin's lymphoma |
| 70842 | Follicular non-Hodg mixed sml cleavd & lge cell lymphoma |
| 99067 | Gamma heavy chain disease |
| 39629 | Granulocytic sarcoma |
| 27330 | Leukaemic reticuloendotheliosis |
| 53397 | Hodgkin's disease NOS |
| 107804 | Hodgkin's disease NOS of intra-abdominal lymph nodes |
| 59755 | Hodgkin's disease NOS of intrathoracic lymph nodes |
| 91900 | Hodgkin's disease NOS of lymph nodes of axilla and arm |
| 59778 | Hodgkin's disease NOS of lymph nodes of head, face and neck |
| 99012 | Hodgkin's disease NOS of lymph nodes inguinal region and leg |
| 97746 | Hodgkin's disease NOS of lymph nodes of multiple sites |
| 94279 | Hodgkin's disease NOS of spleen |
| 61662 | Hodgkin's disease NOS, unspecified site |
| 67703 | Hodgkin's disease, lymphocytic depletion |
| 101530 | Hodgkin's disease, lymphocytic depletion NOS |
| 63625 | Hodgkin's lymphocytic depletion lymph nodes axilla and arm |
| 111942 | Hodgkin's lymphocytic depletion of head, face and neck |
| 110563 | Hodgkin's lymphocytic depletion lymph nodes inguinal and leg |
| 107032 | Hodgkin's lymphocytic depletion lymph nodes multiple sites |
| 101715 | Hodgkin's disease, lymphocytic depletion of spleen |
| 73532 | Hodgkin's, lymphocytic-histiocytic pred intra-abdominal node |
| 95338 | Hodgkin's, lymphocytic-histiocytic pred intrapelvic nodes |
| 92245 | Hodgkin's, lymphocytic-histiocytic pred intrathoracic nodes |
| 68330 | Hodgkin's, lymphocytic-histiocytic pred of head, face, neck |
| 93951 | Hodgkin's, lymphocytic-histiocytic pred inguinal and leg |
| 104743 | Hodgkin's, lymphocytic-histiocytic pred of multiple sites |
| 71142 | Hodgkin's, lymphocytic-histiocytic predominance unspec site |
| 49605 | Hodgkin's disease, mixed cellularity |
| 94005 | Hodgkin's disease, mixed cellularity NOS |
| 58684 | Hodgkin's mixed cellularity of intrathoracic lymph nodes |
| 94407 | Hodgkin's mixed cellularity of lymph nodes head, face, neck |
| 108886 | Hodgkin's mixed cellularity of lymph nodes inguinal and leg |
| 97863 | Hodgkin's disease, mixed cellularity of unspecified site |
| 29178 | Hodgkin's disease, nodular sclerosis |
| 63054 | Hodgkin's disease, nodular sclerosis NOS |
| 61149 | Hodgkin's nodular sclerosis of intra-abdominal lymph nodes |
| 67506 | Hodgkin's nodular sclerosis of intrathoracic lymph nodes |
| 65483 | Hodgkin's nodular sclerosis of lymph nodes of axilla and arm |
| 55303 | Hodgkin's nodular sclerosis of head, face and neck |
| 19140 | Hodgkin's nodular sclerosis of lymph nodes of multiple sites |
| 105472 | Hodgkin's disease, nodular sclerosis of spleen |
| 57225 | Hodgkin's disease, nodular sclerosis of unspecified site |
| 95049 | Hodgkin's lymphocytic depletion of unspecified site |
| 64036 | Hodgkin's sarcoma |
| 68039 | Hodgkin's sarcoma of lymph nodes of axilla and upper limb |
| 29876 | Hodgkin's, lymphocytic-histiocytic predominance NOS |
| 106911 | Hodgkin's, lymphocytic-histiocytic predominance of spleen |
| 43450 | Immunoproliferative neoplasm or myeloma NOS |
| 34926 | Letterer-Siwe disease |
| 47204 | Letterer-Siwe disease NOS |
| 102158 | Letterer-Siwe disease of intrathoracic lymph nodes |
| 54083 | Letterer-Siwe disease of lymph nodes of multiple sites |
| 102715 | Letterer-Siwe disease of unspecified sites |
| 4250 | Leukaemia NOS |
| 25191 | Leukaemia of unspecified cell type |
| 73777 | Leukaemic reticuloendotheliosis NOS |
| 65123 | Leukaemic reticuloend of intra-abdominal lymph nodes |
| 65122 | Leukaemic reticuloendotheliosis of unspecified sites |
| 38939 | Hodgkin's disease, lymphocytic-histiocytic predominance |
| 19372 | Lymphoid leukaemia |
| 38914 | Lymphoid leukaemia NOS |
| 72197 | Lymphosarcoma cell leukaemia |
| 16527 | Macroglobulinaemia |
| 71994 | Macroglobulinaemia NOS |
| 50695 | Diffuse non-Hodgkin mixed sml & lge cell (diffuse) lymphoma |
| 108182 | Diffuse non-Hodgkin's small cleaved cell (diffuse) lymphoma |
| 12335 | Malignant lymphoma NOS |
| 42579 | Malignant lymphoma NOS of intra-abdominal lymph nodes |
| 71262 | Malignant lymphoma NOS of intrapelvic lymph nodes |
| 72725 | Malignant lymphoma NOS of intrathoracic lymph nodes |
| 34089 | Malignant lymphoma NOS of lymph nodes of axilla and arm |
| 50696 | Malignant lymphoma NOS of lymph nodes of head, face and neck |
| 63105 | Malignant lymphoma NOS of lymph node inguinal region and leg |
| 15504 | Malignant lymphoma NOS of lymph nodes of multiple sites |
| 60092 | Malignant lymphoma NOS of spleen |
| 57427 | Malignant lymphoma NOS of unspecified site |
| 17887 | Malignant lymphoma otherwise specified |
| 89657 | Malignant mast cell tumour NOS |
| 15036 | Malignant mast cell tumours |
| 49301 | Malignant neoplasm lymphatic or haematopoietic tissue NOS |
| 30646 | Malignant neoplasm lymphatic or haematopoietic tissue OS |
| 65434 | Malignant neoplasms of lymphoid and histiocytic tissue NOS |
| 22158 | Malignant plasma cell neoplasm, extramedullary plasmacytoma |
| 65721 | Mast cell leukaemia |
| 100615 | Mast cell malignancy of lymph nodes inguinal region and leg |
| 31324 | Mast cell malignancy of lymph nodes of multiple sites |
| 103900 | Mast cell malignancy of unspecified site |
| 57671 | Megakaryocytic leukaemia |
| 15883 | Monoclonal paraproteinaemia |
| 35875 | Monocytic leukaemia |
| 93342 | Monocytic leukaemia NOS |
| 37182 | Multiple myeloma and immunoproliferative neoplasms |
| 12006 | Mycosis fungoides |
| 38005 | Mycosis fungoides NOS |
| 91674 | Mycosis fungoides of intra-abdominal lymph nodes |
| 96379 | Mycosis fungoides of lymph nodes of axilla and upper limb |
| 72714 | Mycosis fungoides of lymph nodes of inguinal region and leg |
| 95012 | Mycosis fungoides of lymph nodes of multiple sites |
| 95949 | Mycosis fungoides of unspecified site |
| 7176 | Myeloid leukaemia |
| 33344 | Myeloid leukaemia NOS |
| 70724 | Myeloid sarcoma |
| 4944 | Multiple myeloma |
| 20440 | Myelomonocytic leukaemia |
| 25493 | Neoplasm of uncertain behaviour of spleen |
| 65701 | Nodular lymphoma NOS |
| 92068 | Nodular lymphoma of intra-abdominal lymph nodes |
| 105203 | Nodular lymphoma of intrathoracic lymph nodes |
| 111766 | Nodular lymphoma of lymph nodes of axilla and upper limb |
| 45264 | Nodular lymphoma of lymph nodes of head, face and neck |
| 94995 | Nodular lymphoma of lymph nodes of inguinal region and leg |
| 58082 | Nodular lymphoma of lymph nodes of multiple sites |
| 66327 | Nodular lymphoma of unspecified site |
| 3604 | Non - Hodgkin's lymphoma |
| 94174 | Other and unspecified leukaemia |
| 99413 | Other and unspecified leukaemia NOS |
| 64567 | Other immunoproliferative neoplasms |
| 34692 | Other leukaemia of unspecified cell type |
| 49725 | Other lymphoid leukaemia |
| 38331 | Other lymphoid leukaemia NOS |
| 99015 | Other monocytic leukaemia |
| 103645 | Other monocytic leukaemia NOS |
| 112440 | Other myeloid leukaemia |
| 66089 | Other myeloid leukaemia NOS |
| 3451 | Other paraproteinaemias |
| 37272 | Other specified leukaemia |
| 30632 | Other specified leukaemia NOS |
| 31576 | Other types of follicular non-Hodgkin's lymphoma |
| 12386 | Paraproteinaemia NOS |
| 2481 | Polycythaemia vera |
| 31586 | Prolymphocytic leukaemia |
| 35014 | Sezary's disease |
| 100532 | Sezary's disease NOS |
| 54793 | Subacute leukaemia NOS |
| 72774 | Subacute lymphoid leukaemia |
| 101606 | Subacute monocytic leukaemia |
| 63475 | Subacute myeloid leukaemia |
| 104475 | Subacute myelomonocytic leukaemia |
| 10411 | Waldenstrom's macroglobulinaemia |
| 5019 | Cancer chemotherapy |
| 102559 | Immunosuppressant drug therapy |
| 59610 | Transplant immunosuppression |
| 105256 | Immunosuppressive therapy |
| 31489 | [V]Chemotherapy session for neoplasm |
| 10505 | Asplenia |
| 23770 | Acquired immune deficiency syndrome |
| 58857 | Acute human immunodeficiency virus infection |
| 58859 | Asymptomatic human immunodeficiency virus infection |
| 69766 | HIV infection with persistent generalised lymphadenopathy |
| 70869 | Human immunodeficiency virus with constitutional disease |
| 53636 | Human immunodeficiency virus with neurological disease |
| 70528 | Human immunodeficiency virus with secondary infection |
| 101836 | Human immunodeficiency virus with secondary cancers |
| 47632 | HIV disease result/haematological+immunologic abnorms,NEC |
| 111979 | HIV disease resulting in multiple diseases CE |
| 67575 | HIV disease resulting in unspecified malignant neoplasm |
| 71450 | HIV disease resulting/unspcf infectious+parasitic disease |
| 62891 | Human immunodeficiency virus with other clinical findings |
| 36294 | Acquired human immunodeficiency virus infection syndrome NOS |
| 44303 | Human immunodef virus resulting in other disease |
| 37006 | HIV disease resulting in mycobacterial infection |
| 66368 | HIV disease resulting in cytomegaloviral disease |
| 23951 | HIV disease resulting in candidiasis |
| 27641 | HIV disease resulting in Pneumocystis carinii pneumonia |
| 50076 | HIV disease resulting in multiple infections |
| 27853 | HIV disease resulting in Kaposi's sarcoma |
| 44617 | HIV disease resulting in Burkitt's lymphoma |
| 66367 | HIV dis resulting oth types of non-Hodgkin's lymphoma |
| 105324 | HIV disease resulting in multiple malignant neoplasms |
| 65117 | HIV disease resulting in lymphoid interstitial pneumonitis |
| 8281 | HIV disease resulting in wasting syndrome |
| 51708 | HIV dis reslt/oth mal neopl/lymph,h'matopoetc+reltd tissu |
| 62854 | [X]Human immunodeficiency virus disease |
| 112030 | [X]HIV disease resulting in other bacterial infections |
| 107807 | [X]HIV disease resulting in other viral infections |
| 112031 | [X]HIV disease resulting in other mycoses |
| 102117 | [X]HIV disease resulting in multiple infections |
| 104134 | [X]HIV disease resulting/other infectious+parasitic diseases |
| 112032 | [X]HIV disease resulting/unspcf infectious+parasitic disease |
| 69767 | [X]HIV disease resulting in other non-Hodgkin's lymphoma |
| 112034 | [X]HIV dis reslt/oth mal neopl/lymph,h'matopoetc+reltd tissu |
| 112035 | [X]HIV disease resulting in other malignant neoplasms |
| 112036 | [X]HIV disease resulting in unspecified malignant neoplasm |
| 112037 | [X]HIV disease resulting in multiple diseases CE |
| 96751 | [X]HIV disease result/haematological+immunologic abnorms,NEC |
| 102252 | [X]HIV disease resulting in other specified conditions |
| 100769 | [X]Unspecified human immunodeficiency virus [HIV] disease |
| 41185 | [X]Dementia in human immunodef virus [HIV] disease |

#### **Supplementary table 3: Medcodes, immcodes, and prodcodes to define Zostavax exposure in CPRD Aurum**

| Immtype | Medcodeid | Prodcodeid | Read Term |
| --- | --- | --- | --- |
| 88 | .. | .. | SHINGLES |
| 91 | .. | .. | SHINGLESOHP |
| .. | 2229311000000111 | .. | Shingles vaccination |
| .. | 1866901000006114 | .. | Herpes zoster vaccination given by other health care provide |
| .. | 1841571000006112 | .. | Herpes zoster vaccination |
| .. | 1340531000033118 | .. | Zostavax |
| .. | 2223901000000112 | .. | Herpes zoster vaccination |
| .. | .. | 6539641000033116 | Zostavax vaccine powder and solvent for suspension for injection 0.65ml pre-filled syringes |
| .. | .. | 6539441000033118 | Shingles (Herpes Zoster) vaccine (live) powder and solvent for suspension for injection 0.65ml pre-filled syringes |
| .. | .. | 2796041000033114 | Varicella vaccine (live) powder and solvent for solution for injection 0.5ml vials |
| .. | .. | 3270541000033113 | Varicella vaccine (live) powder and solvent for suspension for injection 0.5ml vials |

#### **Supplementary table 4: International Classification of Disease and Health Related Problems Revision 10 (ICD-10) code used to define infection-associated hospitalisation (from Torisson et al, Lancet Regional Health Volume 16, 100343, May 2022)**^2^

| **Major ID category Codes** | **ICD10** |
| --- | --- |
| Enteric infections | A00, A01, A02, A03, A04, A05, A06, A07, A08, A09 |
| Bloodstream,infections | A40, A41, R572, R650, R651 |
| Sexually transmitted infections | A50, A51, A52, A53, A54, A55, A56, A57, A58, A59, A60, A61, A62, A63, A64, B20, B21, B22, B23, B24 |
| Infections of the neurological system including eye | A39, A80, A81, A82, A83, A84, A85, A86, A87, A88, A89, B30, G00, G01, G02, G039, G040, G041, G042, G049, G05, G06, G07, G08, G940, H000, H03, H043, H050, H061, H100, H102, H103, H109, H130, H131, H160, H162, H168, H169, H190, H191, H192, H220, H320, H440, H451 |
| URTI including ear | H600, H601, H602, H603, H608, H609, H610, H62, H66, H67, H70, H730, H750, H830, H940, J00, J01, J02, J03, J04, J05, J06, J340, J36, J390, J391 |
| LRTI including influenza | A15, A16, A17, A18, A19, A481,J09, J10, J11, J12, J13, J14, J15, J16, J17, J18, J19, J20, J21, J22, J440, J441, J690, J85, J86 |
| Infectious of the heart and blood vessels | I301, I320, I321, I33, I38, I39, I400, I410, I1411, I412, I430, I520, I521, I681, I790, I791, I980, I981 |
| Infections of the digestive system including liver | B15, B16, B17, B18, B19,K044, K046, K047, K102, K113, K122, K230, K231, K61, K630, K650, K67, K750, K770, K830, K930, K931 |
| Infectious of the GUM system | N080, N10, N136, N151, N159, N160, N290, N291, N300, N309, N330, N340, N341, N37, N390, N410, N412, N413, N431, N450, N459, N481, N482, N49, N51, N61, N70, N71, N72, N73, N74, N751, N764, N770, N771 |
| Infections of the skin and soft tissue | A46, L00, L01, L02, L03, L04, L050, L08, L303 |
| Infections of the bone joints and connective tissue | M00, M01, M462, M463, M464, M465, M490, M491, M492, M600, M630, M631, M632, M650, M651, M680, M710, M711, M726, M730, M731, M86, M900, M901, M902 |
| Infectious complications | T793, T802, T814, T826*, T827*, T835*, T836*, T845, T846, T847*, T857*, T874, T880 |
| Other infections | A20, A21, A22, A23, A24, A25, A26, A27, A28, A30, A31, A32, A33, A34, A35, A36, A37, A38, A42, A43, A44, A45, A46, A47, A48, A49, A65, A66, A67, A68, A69, A70, A71, A72, A73, A74, A75, A76, A77, A78, A79, A92, A93, A94, A95, A96, A97, A98, A99, B00, B01, B02, B03, B04, B05, B06, B07, B08, B09, B25, B26, B27, B33, B34, B35, B36, B37, B38, B39, B40, B41, B42, B43, B44, B45, B46, B47, B48, B49,B50, B51, B52, B53, B54, B55, B56, B57, B58, B59, B60, B61, B62, B63, B64, B65, B66, B67, B68, B69, B70, B71, B72, B73, B74, B75, B76, B77, B78, B79, B80, B81, B82, B83, B84, B85, B86, B87, B88, B89, B90, B91, B92, B93, B94, B95, B96, B97, B98, B99, R508, R509, D709, D733, E060, E32 |

#### **Supplementary table 5: Medcodes, immcodes, and prodcodes to define Varivax exposure in CPRD Aurum**

| Immtype | Medcodeid | Prodcodeid | Read Term |
| --- | --- | --- | --- |
| 43 | .. | .. | VARICELLA |
| .. | 942631000033114 | .. | Varivax |
| .. | 1566281000006112 | .. | 1st varicella vaccination |
| .. | 2695409011 | .. | Second varicella vaccination |
| .. | 1566291000006110 | .. | 2nd varicella vaccination |
| .. | 883421000033118 | .. | Live Attenuated Varicella-Zoster (Oka/Merck Strain) Virus |
| .. | 2674513017 | .. | First varicella vaccination |
| .. | .. | 3187941000033110 | Varivax vaccine powder and solvent for suspension for injection 0.5ml vials (Merck Sharp & Dohme Ltd) |

#### **Supplementary figure 1: Cohort study flowchart of participant eligibility.**

Participants excluded (n=129884):

- n=0 received Varivax
- n=15 had no event date associated with Zostavax dose
- n=1609 received Zostavax dose before current registration date plus 28 weeks
- n=1574 received Zostavax prior to September 2013
- n=42933 died or transferred out of the GP practice prior to entry date (age 69.5 or 1^st^ September 2013)
- n=435 with Zostavax prior to entry date
- n=3060 received Zostavax prior to age 70
- n=13398 with immunosuppression
- n= 64140 receiving PPV23 after entry
- n= 2720 receiving multiple doses of Zostavax
- n=0 outcome occurring more than 30 days after transfer out of practice
- n=0 outcome occurring after practice last-collection date
- n=0 outcome prior to entry date

Records identified from CPRD Aurum. Participants born between 1933 and 1948 who received PPV23.

N=444 502

Cohort for study

N=314 618

Zostavax exposed

N=174534

Zostavax unexposed

N=140084

### **2. STUDY DESIGN**

#### **Supplementary figure 2: Study design for time-varying vaccine exposure and time-to-first and -subsequent event modelling**

A. Cox proportional hazards model with time-varying exposure
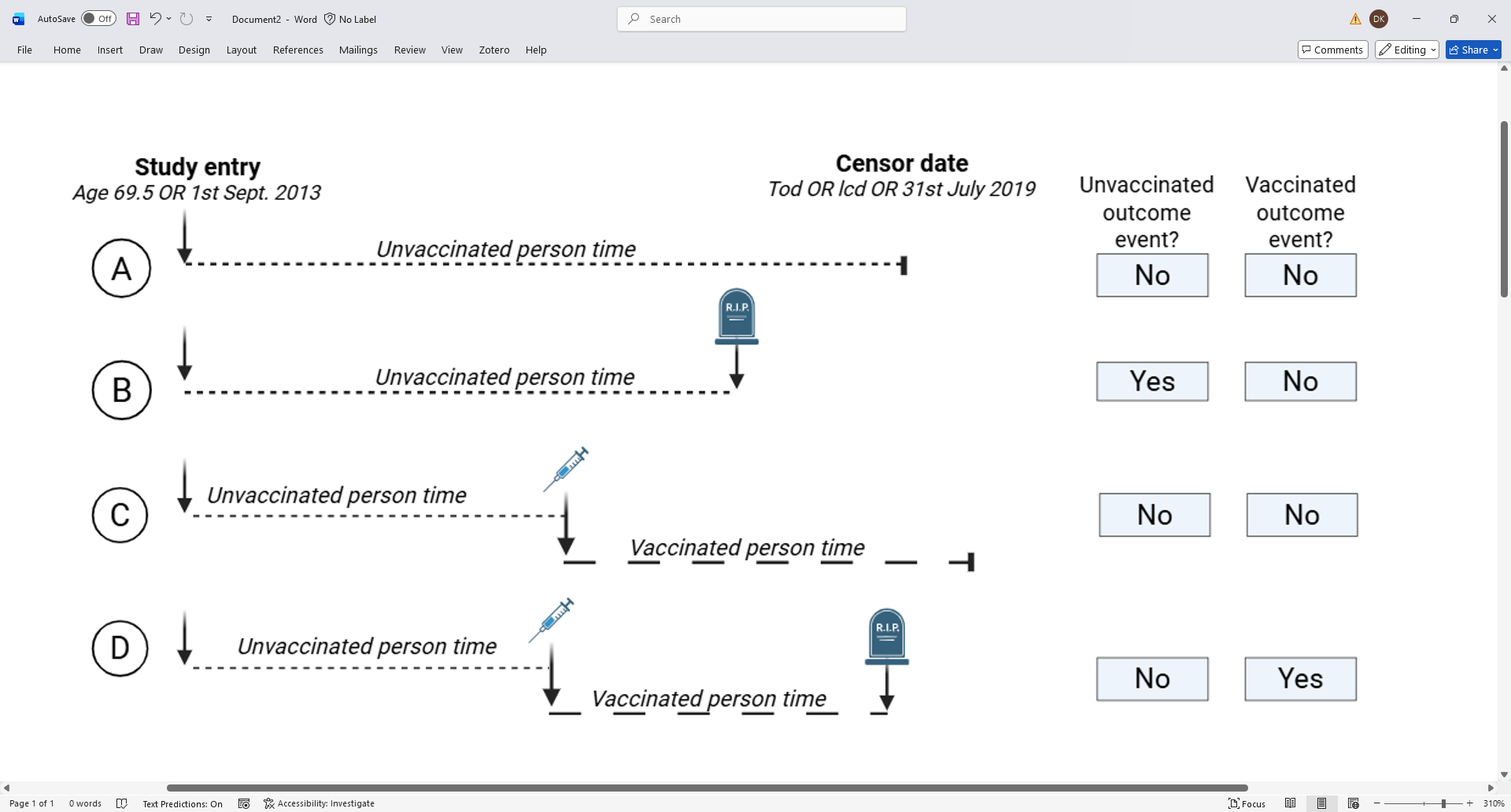

B. Cox-Anderson-Gill model with time-varying exposure

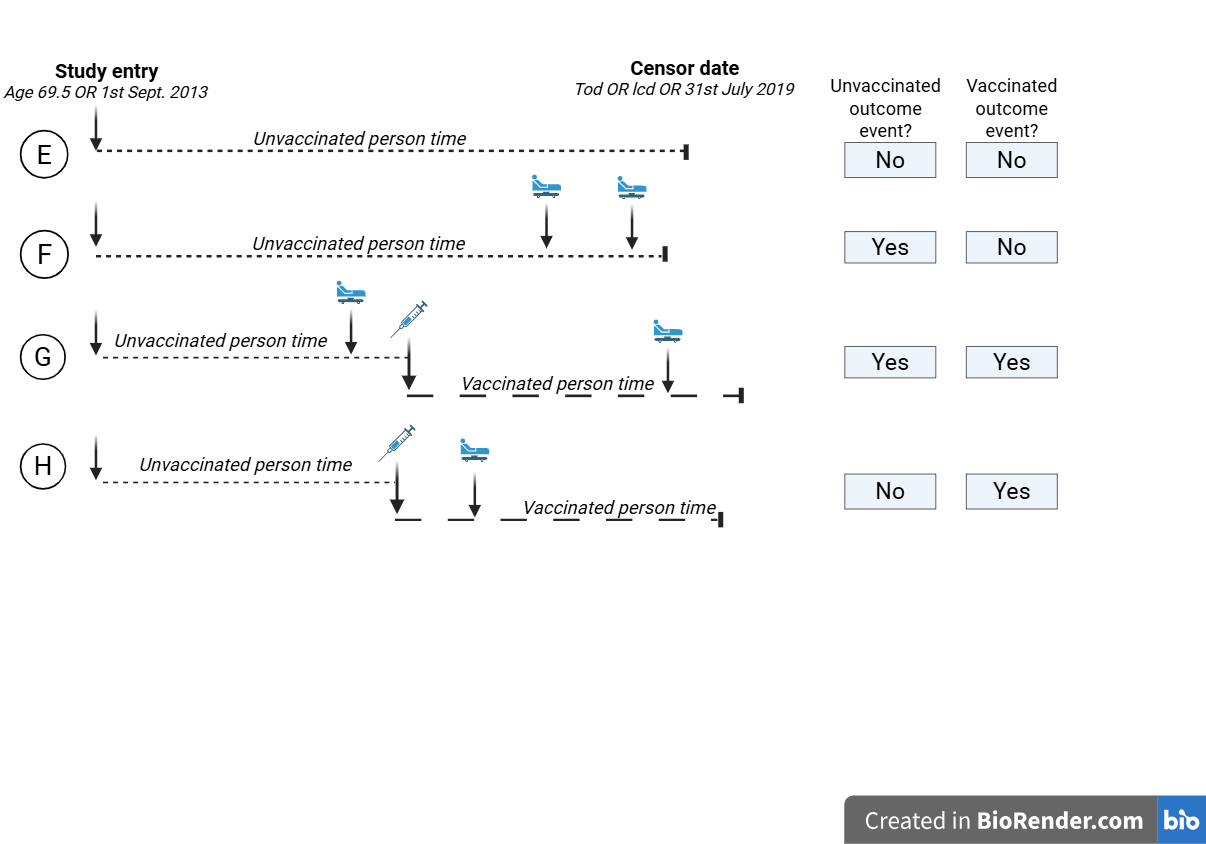

Supplementary figure 2: Study design for time-varying vaccine exposure and time-to-first and -subsequent event modelling. All participants (A-H) enter the study on 1^st^ September 2013 or when they turn 69.5 (whichever is later) and are censored (i.e. follow-up ceases) at transfer-out date (tod) when the individual stops contributing data (due to death or moving medical practice), at last-collection date (lcd) when the medical practice stops contributing quality-assured data, or on 30^th^ June 2019 (whichever is earliest). Panel A: Time-to-first event also censors when an individual experiences an outcome event whether it be death or hospitalisation. Scenario A and B are unvaccinated participants with (B) not experiencing any outcome event and (A) experiencing death during follow-up (
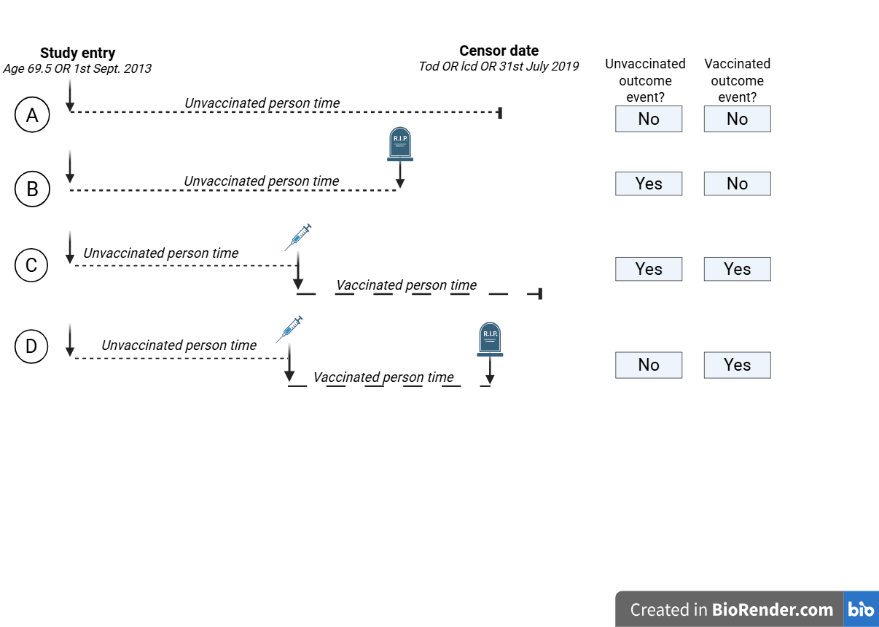
). Scenario C and D are participants who go on to receive live shingles vaccination (
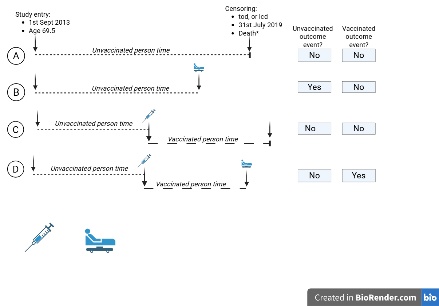
) and contribute both unvaccinated (
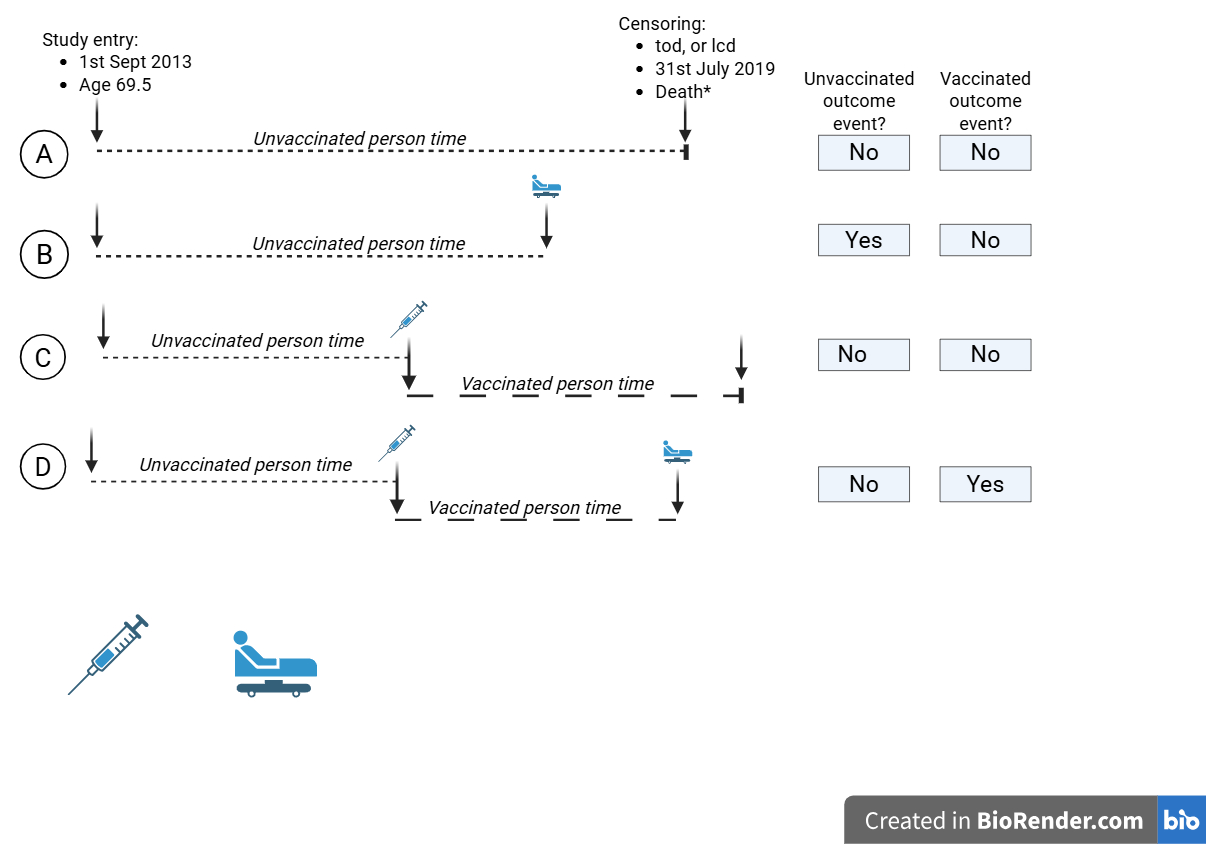
) and vaccinated (
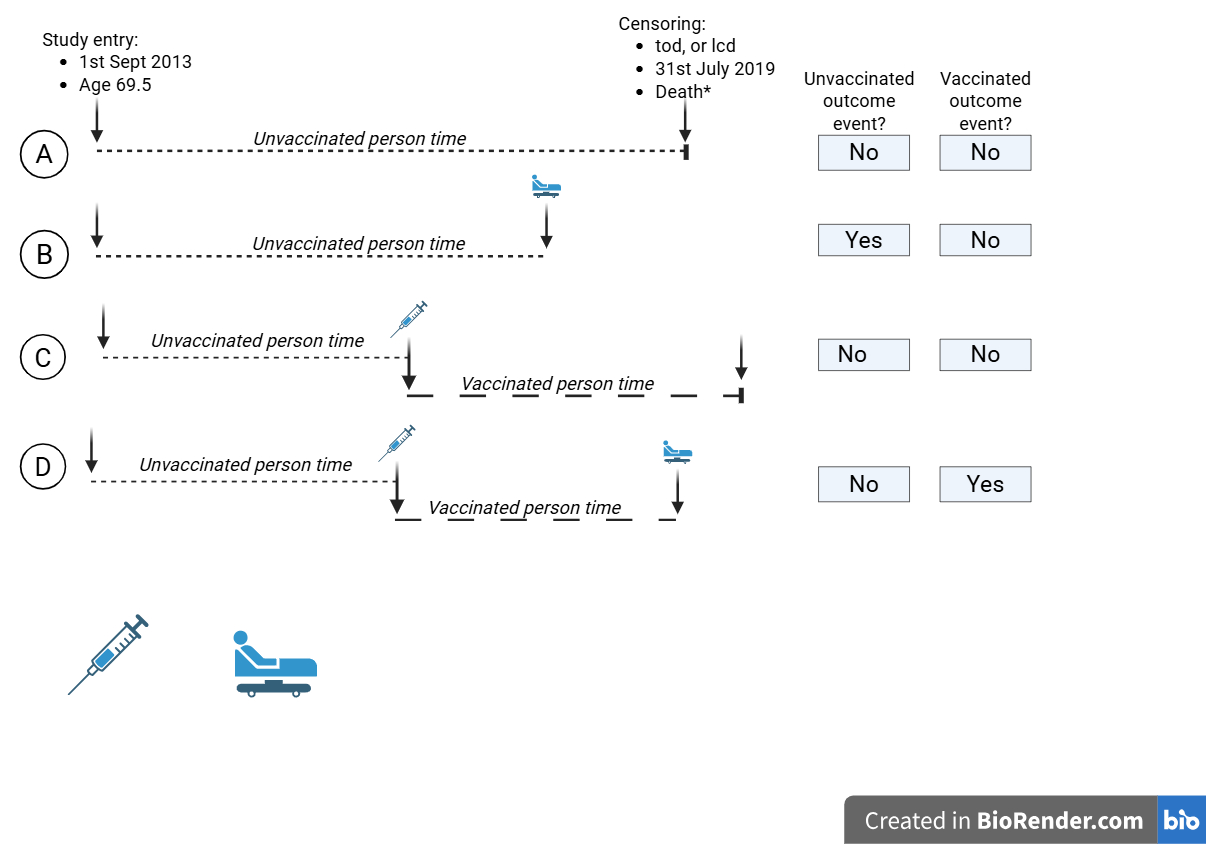
) person-time with (C) having no outcome event and (D) experiencing an outcome event (
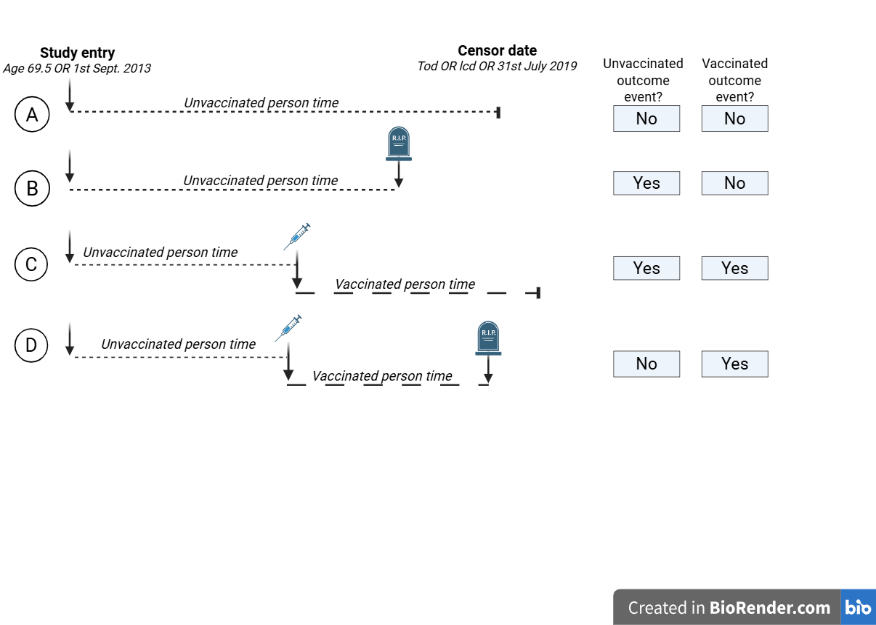
) during vaccinated person-time. Time zero for unvaccinated person-time is study entry and time zero for vaccinated person time is date of vaccination. Panel B: Time-to-subsequent-event modelling in which participants continue contributing person-time after an outcome event. Scenario E and F are unvaccinated participants with (E) not experiencing any outcome event and (F) experiencing two hospitalisation events (
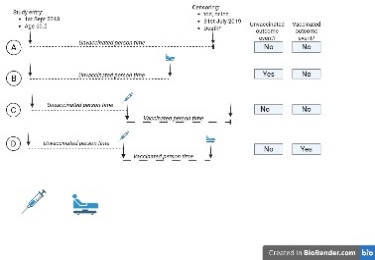
) during follow-up. Scenario G and H are participants who go on to receive live shingles vaccination (
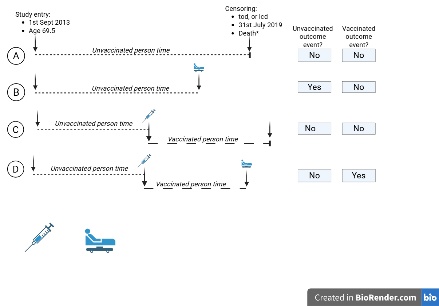
) and contribute both unvaccinated (
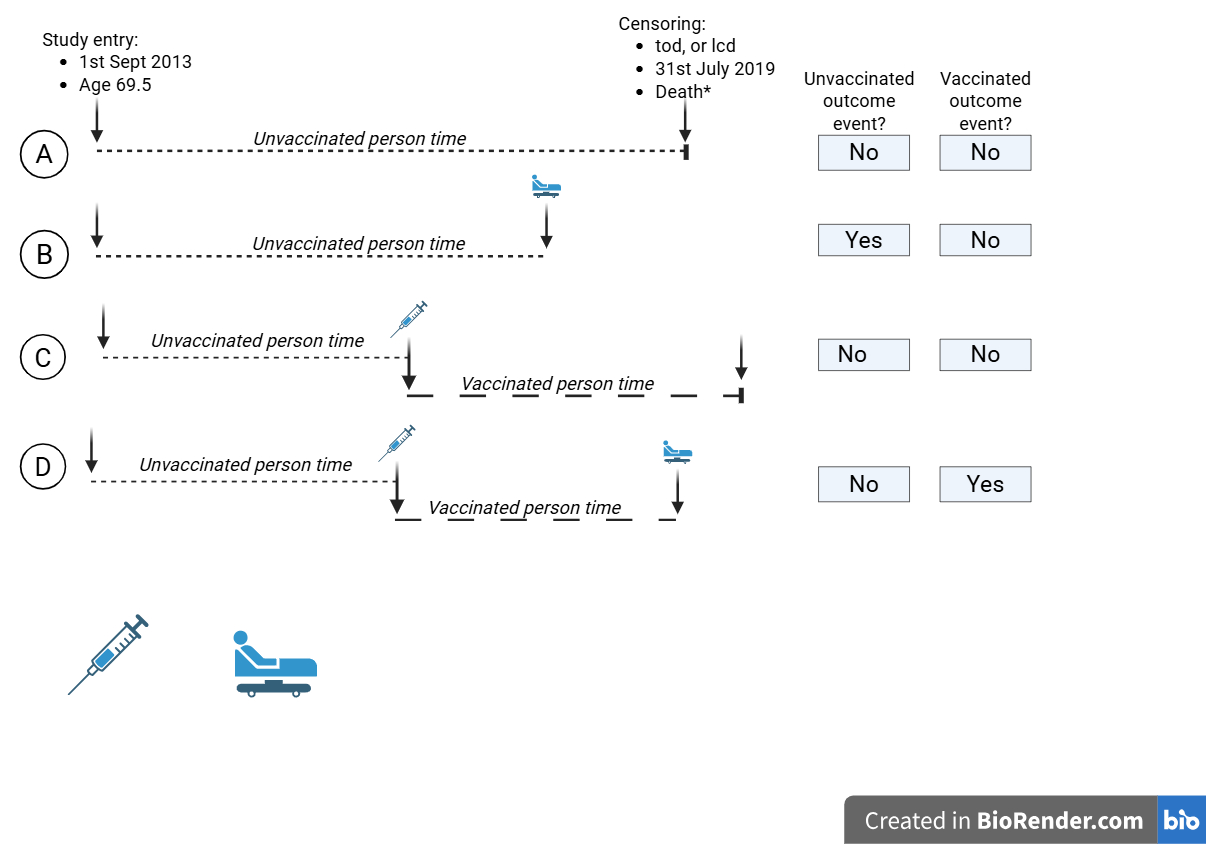
) and vaccinated (
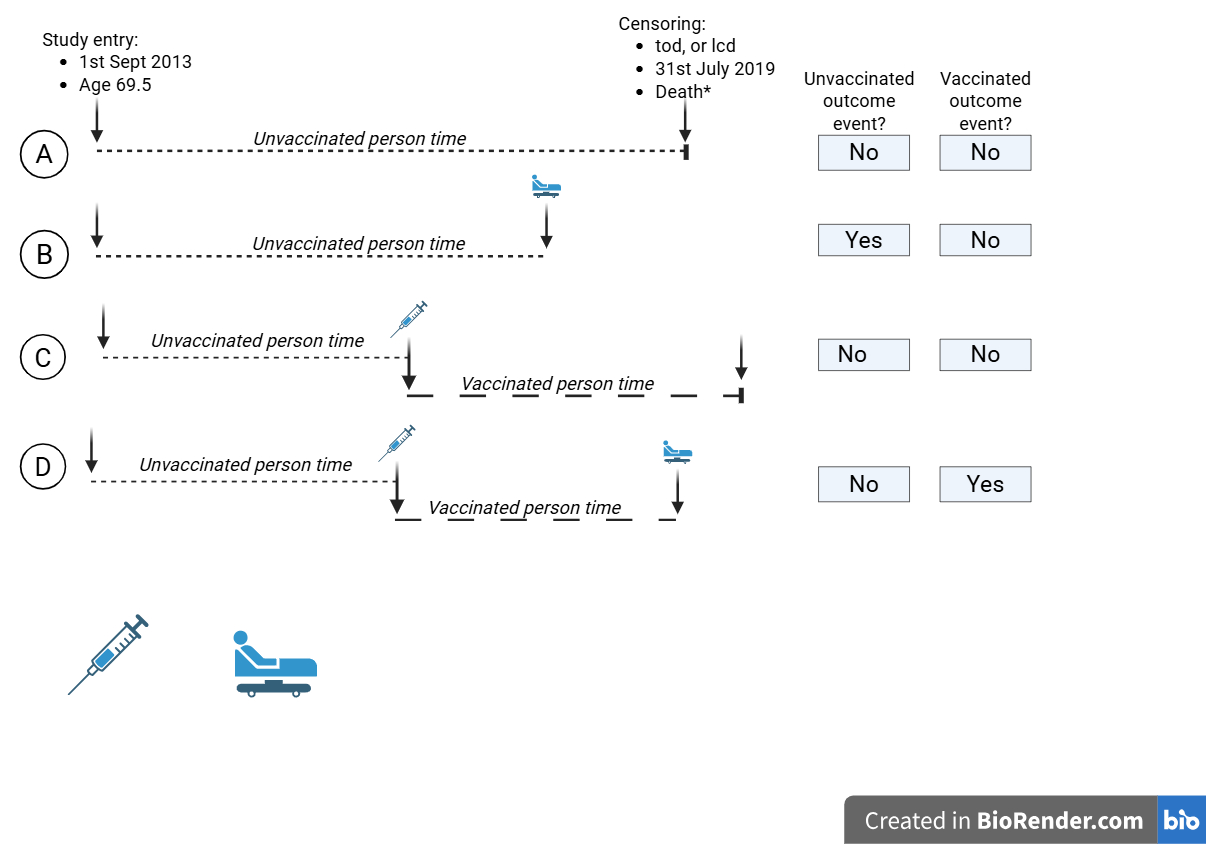
) person-time with (G) having one hospitalisation event (
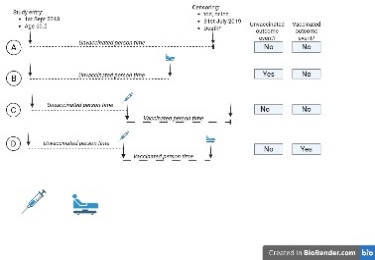
) during unvaccinated person-time and one during vaccinated person time, and (H) experiencing one hospitalisation event (
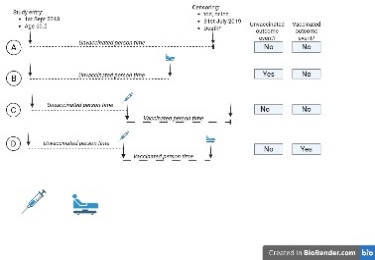
) during vaccinated person-time. Disease rates are reported as number of outcome events/person-time for vaccinated and unvaccinated person time respectively.

#### **3. SUPPLEMENTARY METHODOLOGY: PROPENSITY SCORE GENERATION**

#### **Confounding variables used to generate propensity score of likelihood of live shingles vaccine exposure**

Participant baseline age and sex at study entry were identified from CPRD. To ensure anonymity CPRD only provides year of birth so all participants were given the birthday 1^st^ July for analysis purpose. Participants’ baseline comorbidity status was represented by the 2011 updated version of the Charlson Comorbidity Index; a continuous scale which predicts mortality in high-income settings.^3^ Presence of individual comorbidities from the Charlson comorbidity index were also included in propensity score generation (ischaemic heart disease, heart failure, peripheral vascular disease, cerebrovascular disease, dementia, chronic respiratory disease, connective tissue disease, peptic ulcer disease, chronic liver disease, dementia mellitus, malignancy). Participants were classified at baseline as having ever smoked or having never smoked as ex-smoking status cannot be accurately defined in CPRD.^4^ Socioeconomic status was defined by the patient-level English IMD 2015 quintile and was included in the model as an ordered categorical variable. If patient-level IMD quintile was not available, then practice-level IMD quintile was substituted. Influenza and pneumococcal disease are major causes of mortality and hospitalisation in the study population.^5^ All participants were immunised against pneumococcus with PPV23 prior to study entry. A continuous baseline influenza vaccination score was calculated using influenza vaccine records from the three years prior to study entry. Distinct influenza vaccinations were defined as vaccine events occurring at least six months apart, to avoid duplicate entries for the same vaccination being treated as separate events. The total number of distinct vaccinations was divided by three to generate a score ranging from 0 to 3, with higher values indicating more consistent influenza vaccination over the baseline period. Finally, a continuous variable was generated to reflect participants’ rate of consultation at their medical practice preceding study-entry as this may affect their likelihood of receiving the live shingles vaccine, and their risk of hospitalisation and death. This was generated by calculating the number of weeks in which a participant had contact with their medical practice in the two years preceding the year of study-entry divided by the total number of weeks (104 weeks).^6^

#### **Supplementary figure 3: Covariate balance and common support of propensity score weighting**

**A. Standardised mean difference of covariates between vaccinated and unvaccinated populations using overlap weighting (OW) and inverse probability weighting (IPWT) compared to unweighted**

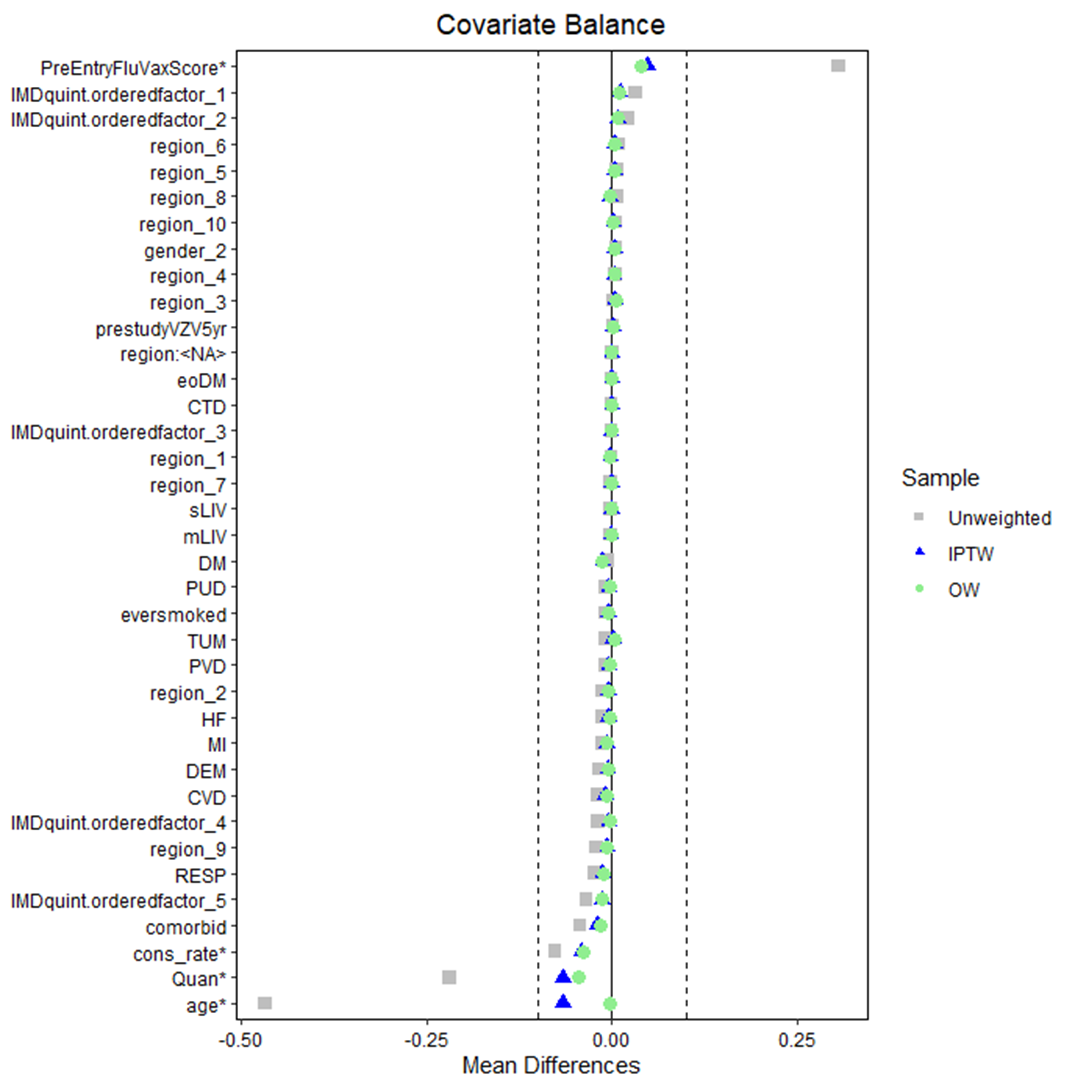

**B. Overlap of propensity score density in vaccinated and unvaccinated populations**

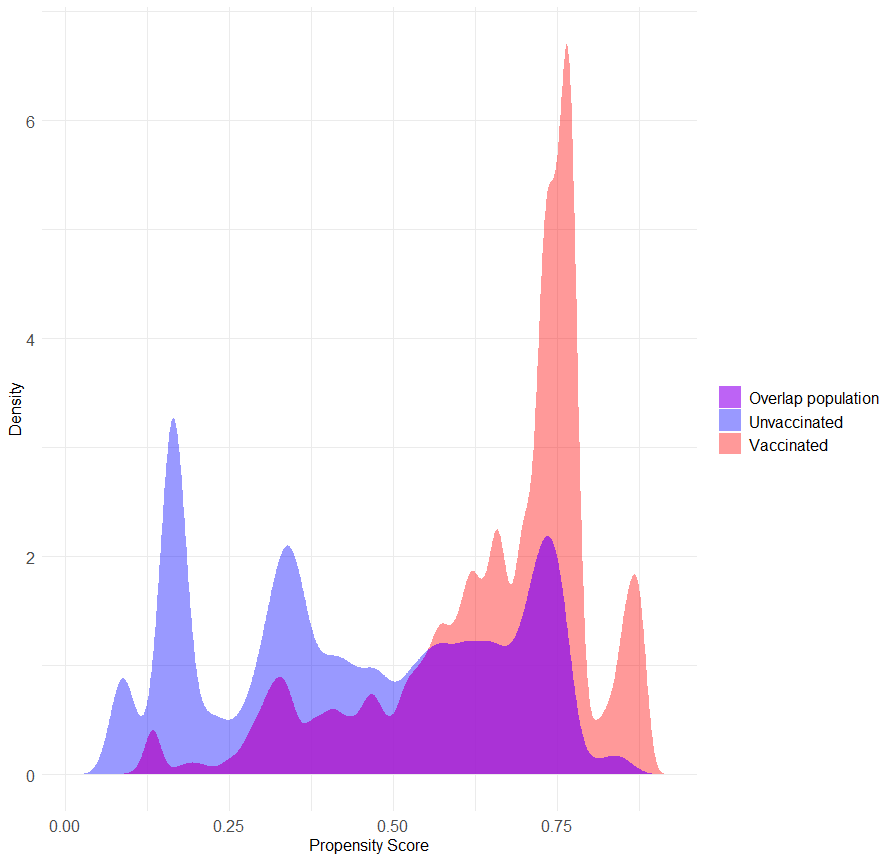

**Supplementary Figure 3. Covariate balance and common support of propensity score weighting.**

(A) Standardised mean differences (SMDs) for baseline covariates comparing vaccinated and unvaccinated individuals before weighting (unweighted) and after adjustment using inverse probability of treatment weighting (IPTW) and overlap weighting (OW). Each point represents the SMD for a covariate; values closer to zero indicate better balance between groups. Vertical dashed lines denote conventional thresholds for acceptable imbalance (SMD < 0.1). Unweighted SMD are not substantially different in the unweighted population likely due to use of a PPV23 vaccinated population. SMD is improved by both IPWT and OW with OW outperforming IPWT for age and comorbidity score (Quan). (B) Distribution of propensity scores in vaccinated and unvaccinated groups, showing the region of common support.

*IPTW=inverse probability of treatment weighting; OW=overlap weighting; SMD=standardised mean difference; Quan = updated Charlson comorbidity index, PUD = peptic ulcer disease; DM=diabetes mellitus; eoDM=diabetes mellitus with end organ damage; CTD=connective tissue disease; sLIV=severe liver disease; mLIV=mild liver disease;TUM=solid organ malignancy; PVD=peripheral vascular disease; HF=heart failure; MI=myocardial ischaemia; DEM=dementia; CVD=cerebrovascular disease; RESP=chronic respiratory disease; cons_rate=rate of consultation at medical practice in two years preceding study entry;IMD=Index of Multiple Deprivation; prestudyVZV5yr=varicella disease in five years preceding entry.*

#### **4. TEST OF PROPORTIONAL HAZARDS**

#### **Supplementary figure 4: Proportion of individuals vaccinated in each year of follow-up**

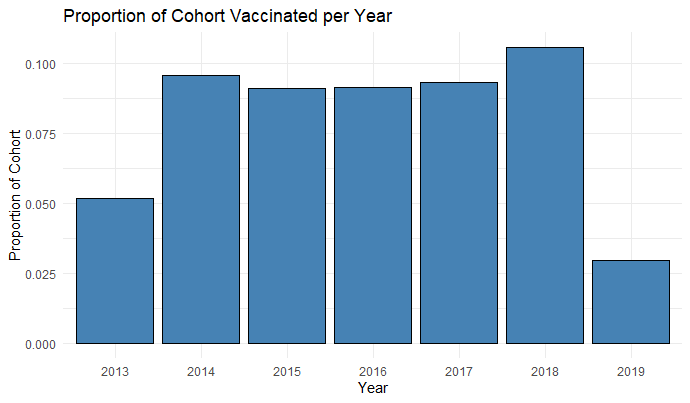

**Supplementary Figure 4. Proportion of individuals vaccinated by calendar year of follow-up.**
Bar chart showing the annual proportion of the study cohort receiving vaccination during each year of follow-up (2013–2019) to interrogate for temporal bias. Bar chart shows stable vaccine coverage by year with lower coverage in 2013 and 2019 reflecting shorter observation time in these years.

#### **Supplementary figure 5: Tests for proportional hazards assumptions for Cox regression models**

**A. Schoenfeld residuals for death:**
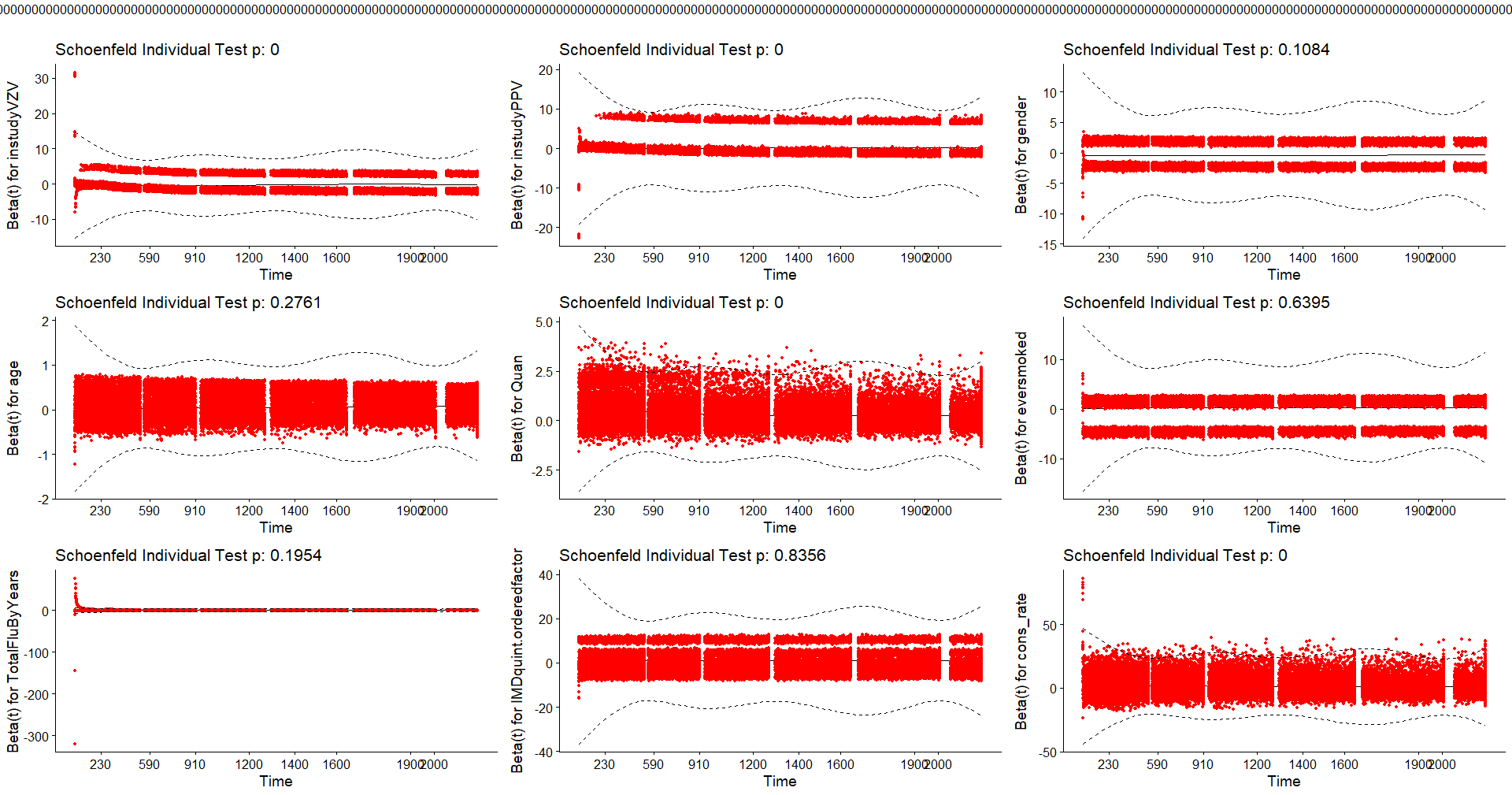

**B. Schoenfeld residuals for hospitalisation:**

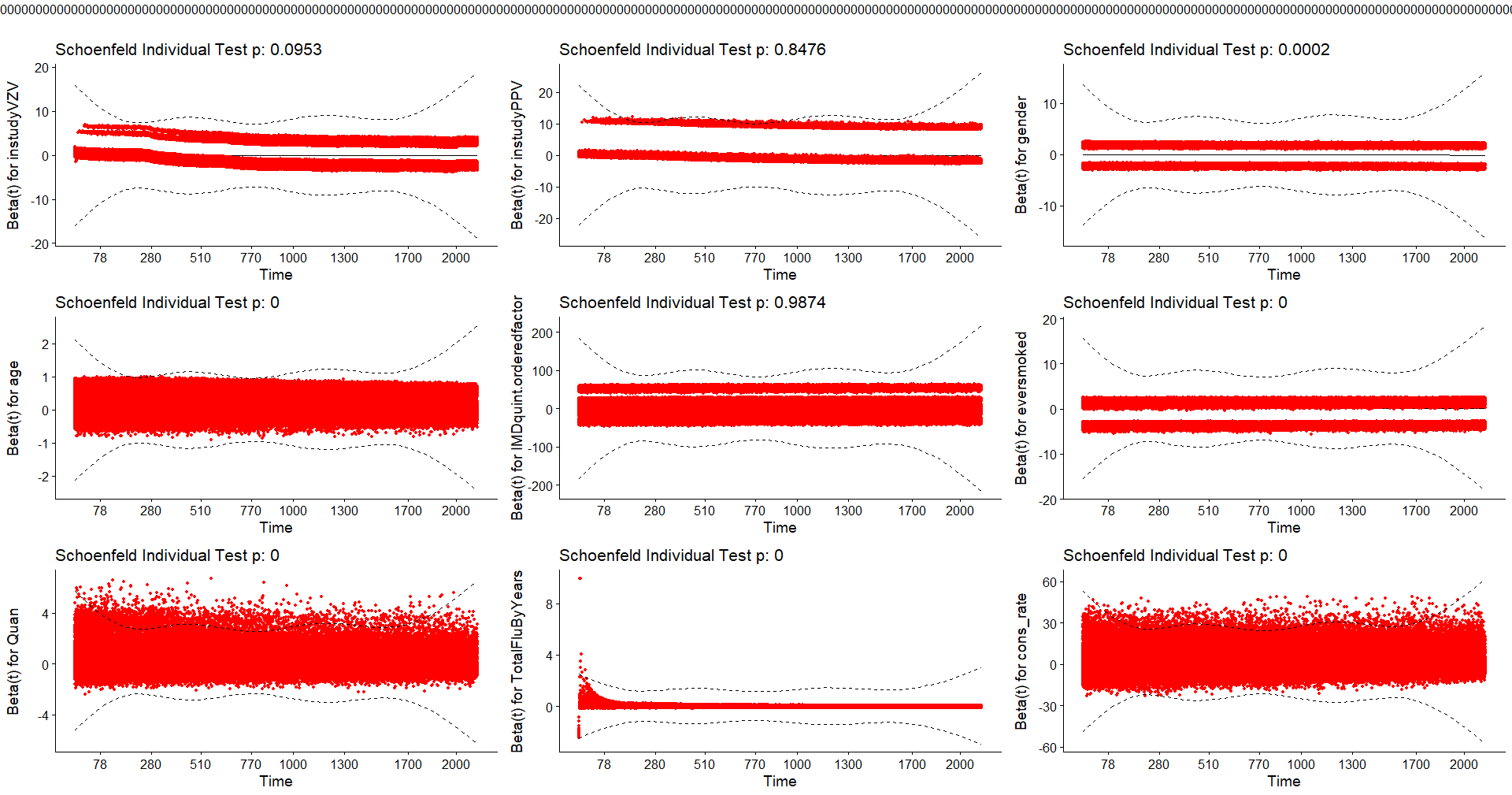

**C. Log-log survival curves for death:**
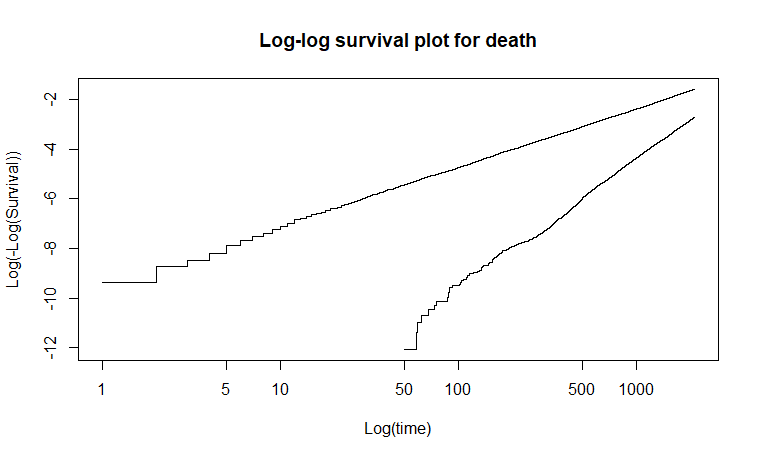

**D. Log-log survival curves for all-cause hospitalisation:**

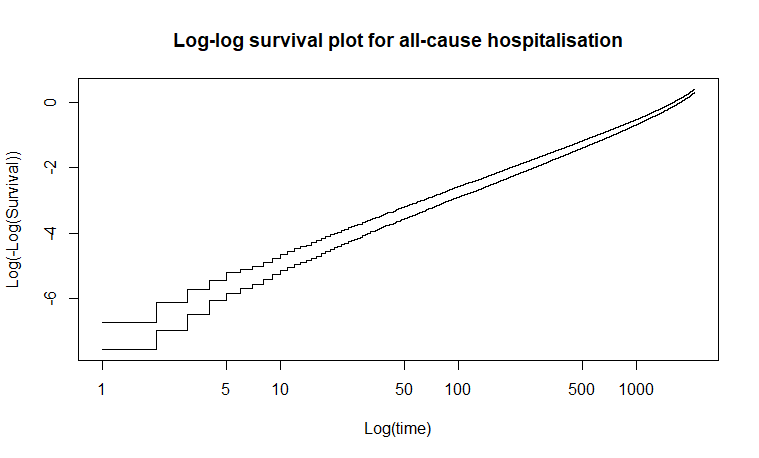

**Supplementary Figure 5. Tests for proportional hazards assumptions for Cox regression models.**
(A–B) Schoenfeld residual plots for covariates for (A) all-cause mortality and (B) all-cause hospitalisation. Scaled Schoenfeld residuals are plotted against follow-up time showing no systematic time trend
(C–D) Log–log survival plots showing line for vaccinated and unvaccinated groups for (C) all-cause mortality and (D) all-cause hospitalisation. The complementary log–log of the estimated survival function is plotted against log(time). Time-zero for unvaccinated person-time was study entry and for vaccinated person-time was date of vaccination. Approximately parallel curves between groups indicate that the proportional hazards assumption is satisfied.

*PH = proportional hazards.*

### **5. SUPPLEMENTARY METHODOLOGY: SUPPLEMENTARY ANALYSES**

#### **Analyses using full source population**

The full source population was used to estimate average treatment/vaccine effect with covariate adjustment for baseline age, sex, Charlson comorbidity Index, Index of Multiple Deprivation quintile, rate of consultation at medical practice, influenza vaccine exposure, and smoking status. In addition the full source population was used to estimate average treatment/vaccine effect with inverse probability weighting using the propensity score described in the main manuscript methods.

#### **Pre-defined sensitivity analyses with removal/addition of sub-populations**

Several pre-defined sensitivity analyses were performed including: Exclusion of participants with less than one year between study entry and censoring who were potentially deemed too frail for live immunisation; inclusion of participants who received the live shingles vaccine prior to the PPV23 vaccine against UK vaccination policy to evaluate potential vaccine sequence effect; exclusion of participants with an outcome event within 30 days of a varicella-zoster disease event to minimise on-target vaccine effects; and exclusion of participants with an outcome event within 14 days of immunisation to minimize the vaccine-associated outcome effects.

#### **Post-hoc statistical analysis: Time-varying coefficient model of hazard ratio over follow-up time**

The large vaccine effect size observed led to the post-hoc decision to evaluate vaccine effect over time following immunisation with the hypothesis that a vaccine effect rather than confounding would be largest immediately after immunisation and attenuate to the null over time. A time-varying coefficient model was used to estimate the hazard ratio over 30-day intervals of follow-up time.^7^ Cubic B-splines were applied to follow-up time with yearly internal knots up to five years and performed better than other spline models including quadratic splines, fewer knot locations, and unspecified knot locations by Akaike's Information Criterion and visualisation plots.^8^ The time-varying coefficient model was run as a univariate Cox proportional hazards model and a multivariate model adjusted for baseline age, sex, smoking status, socioeconomic quintile, influenza vaccine exposure in three years preceding study entry, consultation rate at medical practice in two years preceding entry, and updated Charlson comorbidity score. Computational limits precluded propensity score weighting in the time-varying coefficient model and the random intercept for participants’ medical practice was removed due to convergence failure as a result of low participant numbers in some medical practices.

#### **Absolute measures of survival**

Restricted mean survival time and mean cumulative events were compared in the Cox and Cox-Anderson-Gill models to generate absolute measure of survival.^9,10^

### **6. SUPPLEMENTARY RESULTS: AVERAGE TREATMENT EFFECT USING FULL SOURCE POPULATION WITH COVARIATE ADJUSTMENT OR INVERSE PROBABILITY WEIGHTING**

The hazard ratios were similar in direction and magnitude using the whole original source population with inverse probability weighting or with covariate adjustment compared to the overlap-weighted pseudo-population (Supplementary Tables 7-10).

#### **Supplementary table 6: Baseline characteristics of whole source population aged 70 years and over between September 2013 and June 2019 who have received PPV23 prior to study entry**

|  | Live shingles vaccine unexposed | Live shingles vaccine exposed |
| --- | --- | --- |
| n | 140084 | 174534 |
| Female sex (n (%)) | 73472 (52.4) | 92473 (53.0) |
| Age (mean (SD)) | 72.59 (3.47) | 71.11 (2.81) |
| Comorbid (n (%)) | 89709 (64.0) | 104282 (59.7) |
| Cardiovascular disease | 37680 (26.9) | 23155 (13.3) |
| Respiratory disease | 26706 (19.1) | 29276 (16.8) |
| Liver disease | 1412 (1.0) | 1072 (0.6) |
| Diabetes Mellitus | 33781 (24.1) | 41214 (23.6) |
| Solid organ malignancy | 26850 (19.2) | 32007 (18.3) |
| Dementia | 4116 (2.9) | 2096 (1.2) |
| Connective tissue disease | 2725 (1.9) | 3275 (1.9) |
| Peptic ulcer disease | 6107 (4.4) | 6232 (3.6) |
| Updated Charlson Index of Comorbidity* (mean, SD) | 4.08 (1.40) | 3.81 (1.12) |
| Index Multiple Deprivation quintile** (n (%)) |  |  |
| 1 | 36907 (26.3) | 51520 (29.5) |
| 2 | 31398 (22.4) | 42862 (24.6) |
| 3 | 27919 (19.9) | 34669 (19.9) |
| 4 | 23858 (17.0) | 26472 (15.2) |
| 5 | 20002 (14.3) | 19011 (10.9) |
| Geographical region (n (%)) |  |  |
| Northeast | 6616 (4.7) | 8087 (4.6) |
| Northwest | 24846 (17.7) | 28911 (16.6) |
| Yorkshire & The Humber | 5572 (4.0) | 7353 (4.2) |
| East Midlands | 3240 (2.3) | 4795 (2.7) |
| West Midlands | 26151 (18.7) | 33743 (19.3) |
| East of England | 8338 (6.0) | 11959 (6.9) |
| Southwest | 20716 (14.8) | 25577 (14.7) |
| South Central | 17579 (12.6) | 22987 (13.2) |
| London | 15861 (11.3) | 16178 (9.3) |
| Southeast Coast | 11146 (8.0) | 14924 (8.6) |
| Ever smoked (n (%)) | 100226 (71.5) | 123479 (70.7) |
| Rate of attendance at doctor’s practice in two years preceding study entry*** (mean, SD) | 0.20 (0.13) | 0.19 (0.12) |
| Number of influenza vaccinations per year in three years preceding study entry**** (mean, SD) | 0.80 (0.34) | 0.89 (0.26) |
| Varicella-Zoster associated disease in five years preceding study entry (n (%)) | 5956 (4.25) | 7975 (4.57) |
| **a continuous scale of comorbidity which predict mortality*^3^*; **n=299 participants with missing index of multiple deprivation, corresponding medical practice’s index of multiple deprivation was substituted; ***number of weeks in which participant attended their medical practice in two years preceding year of entry to study / total of 104 weeks; **** number of influenza immunisation events in three years preceding study entry/ 3 giving score between 0 and 3 SD=standard deviation* | | |

#### **Supplementary table 7: Univariate and multivariate Cox survival analysis**

|  | Vaccine unexposed rate/1000pyo | Vaccine exposed rate/1000pyo | CoxPH | |
| --- | --- | --- | --- | --- |
|  |  |  | HR (95% CI) | aHR (95% CI) |
| Death  #outcomes/person-time years | 23.0  19351/839840 | 14.9  7045/473345 | 0.54 (0.53-0.56) | 0.69  (0.67-0.71) |
| Hospitalisation  #outcomes/person-time years | 216  127880/592808 | 219  61360/279938 | 0.96 (0.95-0.97) | 1.00 (0.99-1.01) |
| Infectious hospitalisation  #outcomes/person-time years | 51.2  40541/777982 | 42.8  18509/432879 | 0.71  (0.71-0.74) | 0.85  (0.84-0.87) |

#### **Supplementary table 8: Univariate and multivariate Cox Andersen Gill recurrent event analysis**

|  | Vaccine unexposed rate/1000pyo | Vaccine exposed rate/1000pyo | Cox-Andersen-Gill | |
| --- | --- | --- | --- | --- |
|  |  |  | HR (95% CI) | aHR (95% CI) |
| Hospitalisation  #outcomes/person-time years | 643  663870/1032802 | 560  330157/589860 | 0.79  (0.77-0.82) | 0.85  (0.82-0.88) |
| Infectious hospitalisation  #outcomes/person-time years | 89.7  92996/1036906 | 69.0  40821/591854 | 0.65  (0.64-0.67) | 0.73  (0.72-0.75) |

#### **Supplementary table 9: Cox survival analysis with inverse probability weighting**

|  | Vaccine unexposed rate/1000pyo | Vaccine exposed rate/1000pyo | Cox PH with IPWT |
| --- | --- | --- | --- |
|  |  |  | HR (95% CI) |
| Death  #outcomes/person-time years | 21.2  16807/792546 | 16.8  7388/440893 | 0.67  (0.65-0.69) |
| Hospitalisation  #outcomes/person-time years | 215  121195/563202 | 225  57625/256613 | 0.98  (0.97-1.00) |
| Infectious hospitalisation  #outcomes/person-time years | 50.1  36918/736841 | 46.1  18476/400550 | 0.82  (0.80-0.84) |

#### **Supplementary table 10: Cox Andersen Gill recurrent events model with inverse probability weighting**

|  | Vaccine unexposed rate/1000pyo | Vaccine exposed rate/1000pyo | Cox-Andersen-Gill with IPWT |
| --- | --- | --- | --- |
|  |  |  | HR (95% CI) |
| Hospitalisation  #outcomes/person-time years | 621  603110/970967 | 588  338754/576422 | 0.86 (0.83-0.89) |
| Infectious hospitalisation  #outcomes/person-time years | 83.7  81603/974685 | 75.2  43504/578475 | 0.77 (0.75-0.78) |

### **7. SUPPLEMENTARY RESULTS: SENSITIVITY ANALYSES WITH REMOVAL/ADDITION OF SUB-POPULATIONS**

#### **Supplementary table 11: Rate of hospitalisation and death per 1000 person-years in exposed and unexposed participants excluding participants with less than one-year follow-up** **(n=11877 excluded)**

|  | No shingles vaccine exposure | Shingles vaccine exposure | Cox PH with overlap-weighting* (HR, 95% CI) |
| --- | --- | --- | --- |
| Death per 1000 pyo | 17.4 | 16.9 | 0.65 (0.63-0.67) |
| Hospitalisation per 1000 pyo | 211 | 228 | 0.99 (0.98-1.00) |
| Inf. Hosp. per 1000 pyo | 48.8 | 47.4 | 0.81 (0.79-0.83) |
| *and a random intercept for practice, pyo=person-years of observation* | | | |

#### **Supplementary table 12: Rate of hospitalisation and death per 1000 person-years in exposed and unexposed participants including participants with pneumococcal polysaccharide vaccination after study entry (****n=63519 included)**

|  | No shingles vaccine exposure | Shingles vaccine exposure | Cox PH with overlap-weighting* (HR, 95% CI) |
| --- | --- | --- | --- |
| Death per 1000 pyo | 21.0 | 16.8 | 0.65 (0.63-0.67) |
| Hospitalisation per 1000 pyo | 215 | 227 | 0.98 (0.97-0.99) |
| Inf. Hosp. per 1000 pyo | 51.3 | 46.9 | 0.80 (0.78-0.82) |
| ** and a random intercept for practice, pyo=person-years of observation* | | | |

#### **Supplementary table 13: Rate of hospitalisation and death per 1000 person-years in exposed and unexposed participants without participants with an outcome event within 14 days of live shingles vaccination (n=907 removed)**

|  | No shingles vaccine exposure | Shingles vaccine exposure | Cox PH with overlap-weighting* (HR, 95% CI) |
| --- | --- | --- | --- |
| Death per 1000 pyo | 23.38 | 17.3 | 0.63 (0.61-0.65) |
| Hospitalisation per 1000 pyo | 219 | 225 | 0.98 (0.96-0.98) |
| Inf. Hosp. per 1000 pyo | 52.3 | 47.2 | 0.80 (0.78-0.81) |
| ** and a random intercept for practice, pyo=person-years of observation* | | | |

#### **Supplementary table 14: Rate of hospitalisation and death per 1000 person-years in exposed and unexposed participants excluding participants with an outcome event within 30 days of a varicella-zoster associated disease (n=1221 removed)**

|  | No shingles vaccine exposure | Shingles vaccine exposure | Cox PH with overlap-weighting* (HR, 95% CI) |
| --- | --- | --- | --- |
| Death per 1000 pyo | 23.1 | 17.4 | 0.64 (0.62-0.66) |
| Hospitalisation per 1000 pyo | 217 | 2228 | 0.99 (0.98-1.01) |
| Inf. Hosp. per 1000 pyo | 51.7 | 47.4 | 0.82 (0.80-0.84) |
| ** and a random intercept for practice, pyo=person-years of observation* | | | |

#### **Supplementary table 15: Rate of hospitalisation and death per 1000 person-years in exposed and unexposed participants excluding those in ‘catch-up’ population (year of birth 1933-1942 who were offered live shingles vaccine aged 71-79 in 2013 during initial roll-out) (n=136079 removed)**

|  | No shingles vaccine exposure | Shingles vaccine exposure | Cox PH with overlap-weighting* (HR, 95% CI) |
| --- | --- | --- | --- |
| Death per 1000 pyo | 20.3 | 11.0 | 0.43 (0.40-0.45) |
| Hospitalisation per 1000 pyo | 215 | 210 | 0.96 (0.94-0.98) |
| Inf. Hosp. per 1000 pyo | 45.8 | 35.9 | 0.69 (0.66-0.72) |
| **and a random intercept for practice, pyo=person-years of observation* | | | |

#### **Supplementary table 16: Rate of death per 1000 person-years in exposed and unexposed participants stratified by sex, comorbidity, index of multiple deprivation, smoking status, influenza vaccine exposure in overlap-weighted pseudo-population.**

|  | No shingles vaccine exposure | Shingles vaccine exposure | Cox PH with overlap-weighting (HR, 95% CI) |
| --- | --- | --- | --- |
| All | 23.3 | 17.5 | 0.64 (0.62-0.66) |
| Female | 18.9 | 13.3 | 0.59 (0.56-0.62) |
| Male | 28.1 | 22.1 | 0.67 (0.65-0.70) |
| Age 70 to 74 years | 18.6 | 11.7 | 0.56 (0.54-0.58) |
| Aged 75 years and over | 39.6 | 34.6 | 0.72 (0.68-0.76) |
| With respiratory disease | 41.9 | 32.2 | 0.66 (0.62-0.70) |
| Without respiratory disease | 19.2 | 14.4 | 0.63 (0.61-0.66) |
| With cardiovascular disease | 49.1 | 42.1 | 0.75 (0.71-0.80) |
| Without cardiovascular dis. | 19.2 | 13.8 | 0.61 (0.59-0.63) |
| With diabetes mellitus | 30.0 | 24.4 | 0.67 (0.63-0.71) |
| Without diabetes mellitus | 21.2  24 | 15.5 | 0.63 (0.60-0.65) |
| With solid organ malignancy | 36.8 | 23.1 | 0.58 (0.55-0.62) |
| Without solid organ malig. | 20.2 | 16.1 | 0.66 (0.63-0.68) |
| Charlson 3 (least comorbid) | 11.2 | 9.31 | 0.63 (0.59-0.67) |
| Charlson 4 | 24.3 | 20.2 | 0.67 (0.62-0.71) |
| Charlson 5 | 33.0 | 20.4 | 0.56 (0.52-0.60) |
| Charlson 6 | 50.9 | 37.2 | 0.65 (0.60-0.71) |
| Charlson 7 | 74.4 | 56.4 | 0.66 (0.59-0.75) |
| Charlson 8 | 88.7 | 84.3 | 0.81 (0.66-1.00) |
| Charlson 9 | 201 | 106 | 0.58 (0.48-0.70) |
| Charlson 10 to 15 (most comorbid) | 256 | 137 | 0.61 (0.46-0.79) |
| IMD 1 (least deprived) | 18.2 | 13.1 | 0.62 (0.58-0.66) |
| IMD 2 | 20.7 | 14.7 | 0.61 (0.56-0.65) |
| IMD 3 | 23.4 | 17.4 | 0.63 (0.58-0.67) |
| IMD 4 | 26.6 | 20.6 | 0.66 (0.61-0.71) |
| IMD 5 (most deprived) | 33.7 | 28.8 | 0.72 (0.67-0.78) |
| Ever smoked | 25.9 | 19.8 | 0.65 (0.63-0.67) |
| Never smoked | 16.6 | 11.6 | 0.60 (0.56-0.64) |
| <=0.5 vax/year | 39.5 | 17.5 | 0.57 (0.51-0.63) |
| >0.5<1.0 vax/year | 16.8 | 14.4 | 0.48 (0.46-0.51) |
| >=1.0 vax/year | 23.4 | 21.0 | 0.85 (0.81-0.89) |

#### **Supplementary table 17: Rate of hospitalisation per 1000 person-years in exposed and unexposed participants stratified by sex, comorbidity, index of multiple deprivation, smoking status, influenza vaccine exposure in overlap-weighted pseudo-population.**

|  | No shingles vaccine exposure | Shingles vaccine exposure | Cox PH with overlap-weighting (HR, 95% CI) |
| --- | --- | --- | --- |
| All | 218 | 228 | 0.99 (0.97-1.00) |
| Female | 211 | 218 | 0.98 (0.96-1.00) |
| Male | 227 | 240 | 0.99 (0.98-1.01) |
| Age 70 to 74 years | 212 | 212 | 0.97 (0.96-0.98) |
| Aged 75 years and over | 243 | 286 | 1.02 (0.99-1.05) |
| With respiratory disease | 288 | 293 | 0.95 (0.93-0.98) |
| Without respiratory disease | 204 | 216 | 0.99 (0.98-1.01) |
| With cardiovascular disease | 319 | 321 | 0.99 (0.96-1.02) |
| Without cardiovascular dis. | 204 | 217 | 0.99 (0.98-1.00) |
| With diabetes mellitus | 252 | 262 | 0.99 (0.97-1.01) |
| Without diabetes mellitus | 208 | 219 | 0.99 (0.97-1.00) |
| With solid organ malignancy | 301 | 285 | 0.97 (0.94-0.99) |
| Without solid organ malig. | 202 | 217 | 0.99 (0.97-1.00) |
| Charlson 3 (least comorbid) | 173 | 193 | 1.00 (0.98-1.02) |
| Charlson 4 | 230 | 249 | 0.98 (0.96-1.01) |
| Charlson 5 | 278 | 271 | 0.96 (0.93-0.99) |
| Charlson 6 | 341 | 328 | 0.96 (0.92-1.00) |
| Charlson 7 | 392 | 369 | 0.95 (0.87-1.03) |
| Charlson 8 | 451 | 392 | 0.86 (0.75-0.99) |
| Charlson 9 | 643 | 448 | 0.90 (0.78-1.04) |
| Charlson 10 to 15 (most comorbid) | 756 | 446 | 0.72 (0.57-0.91) |
| IMD 1 (least deprived) | 205 | 212 | 0.97 (0.95-1.00) |
| IMD 2 | 212 | 221 | 0.98 (0.96-1.00) |
| IMD 3 | 219 | 229 | 0.98 (0.96-1.01) |
| IMD 4 | 227 | 239 | 1.00 (0.97-1.03) |
| IMD 5 (most deprived) | 246 | 266 | 1.03 (1.00-1.06) |
| Ever smoked | 229 | 238 | 0.97 (0.96-0.99) |
| Never smoked | 192 | 206 | 1.02 (0.99-1.04) |
| <=0.5 vax/year | 210 | 206 | 1.02 (0.98-1.07) |
| >0.5<1.0 vax/year | 217 | 221 | 0.93 (0.92-0.95) |
| >=1.0 vax/year | 223 | 243 | 0.99 (0.97-1.01) |

#### **Supplementary table 18: Rate of infection-associated hospitalisation per 1000 person-years in exposed and unexposed participants stratified by sex, comorbidity, index of multiple deprivation, smoking status, influenza vaccine exposure in overlap-weighted pseudo-population.**

|  | No shingles vaccine exposure | Shingles vaccine exposure | Cox PH with overlap-weighting (HR, 95% CI) |
| --- | --- | --- | --- |
| All | 52.3 | 47.6 | 0.81 (0.79-0.83) |
| Female | 49.2 | 43.1 | 0.78 (0.75-0.80) |
| Male | 55.8 | 52.8 | 0.84 (0.81-0.86) |
| Age 70 to 74 years | 45.7 | 37.8 | 0.76 (0.74-0.78) |
| Aged 75 years and over | 76.7 | 79.1 | 0.88 (0.85-0.92) |
| With respiratory disease | 96.6 | 86.2 | 0.81 (0.78-0.84) |
| Without respiratory disease | 43.2 | 40.2 | 0.81 (0.79-0.83) |
| With cardiovascular disease | 99.9 | 92.0 | 0.84 (0.81-0.87) |
| Without cardiovascular dis. | 45.3 | 41.7 | 0.80 (0.78-0.82) |
| With diabetes mellitus | 70.1 | 68.2 | 0.86 (0.83-0.89) |
| Without diabetes mellitus | 47.0 | 42.1 | 0.79 (0.77-0.81) |
| With solid organ malignancy | 67.8 | 56.4 | 0.80 (0.77-0.84) |
| Without solid organ malig. | 48.8 | 45.6 | 0.81 (0.79-0.83) |
| Charlson 3 (least comorbid) | 32.2 | 31.4 | 0.80 (0.78-0.83) |
| Charlson 4 | 64.9 | 61.2 | 0.81 (0.78-0.84) |
| Charlson 5 | 64.1 | 53.9 | 0.79 (0.75-0.83) |
| Charlson 6 | 101 | 88.3 | 0.83 (0.78-0.88) |
| Charlson 7 | 140 | 122 | 0.82 (0.74-0.91) |
| Charlson 8 | 177 | 153 | 0.86 (0.72-1.02) |
| Charlson 9 | 217 | 144 | 0.73 (0.60-0.88) |
| Charlson 10 to 15 (most comorbid) | 267 | 172 | 0.69 (0.52-0.91) |
| IMD 1 (least deprived) | 42.4 | 37.5 | 0.79 (0.75-0.83) |
| IMD 2 | 46.5 | 43.2 | 0.82 (0.79-0.86) |
| IMD 3 | 51.6 | 46.3 | 0.79 (0.75-0.82) |
| IMD 4 | 59.8 | 54.3 | 0.80 (0.77-0.85) |
| IMD 5 (most deprived) | 75.9 | 74.5 | 0.88 (0.84-0.93) |
| Ever smoked | 57.6 | 52.5 | 0.81 (0.79-0.83) |
| Never smoked | 39.2 | 35.9 | 0.81 (0.77-0.85) |
| <=0.5 vax/year | 57.2 | 42.2 | 0.77 (0.72-0.83) |
| >0.5<1.0 vax/year | 49.8 | 43.3 | 0.67 (0.65-0.69) |
| >=1.0 vax/year | 52.8 | 53.9 | 0.94 (0.91-0.97) |

### **8. SUPPLEMENTARY RESULTS: TIME-VARYING COEFFICIENT MODEL OF HAZARD RATIO OVER TIME**

#### **Supplementary Figure 6: Hazard ratio associated with live shingles vaccine exposure over follow-up time**

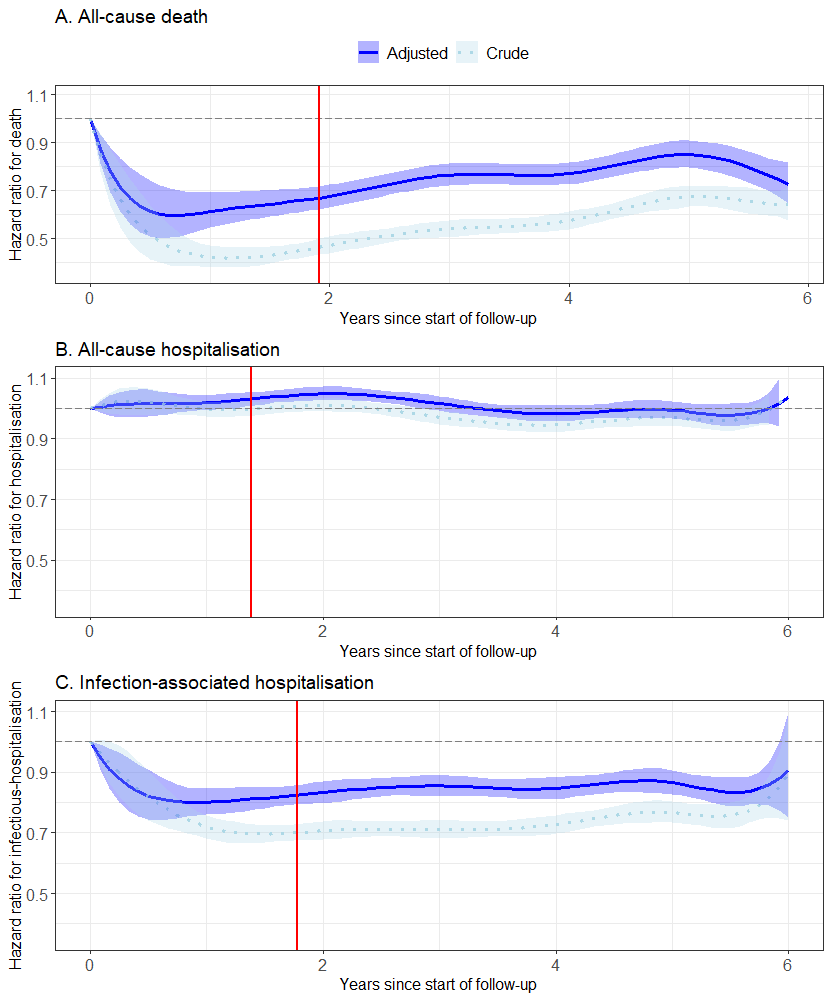

**Figure 2: Time-varying coefficient model of hazard associated with live shingles vaccine exposure over follow-up time.** Hazard ratio for time-to-first event was calculated over 30-day intervals from study entry for unvaccinated person-time and from date of live shingles vaccination for vaccinated person-time. Time-zero for unvaccinated person-time was study entry and for vaccinated person-time was date of vaccination. Cubic B-splines were applied to follow-up time with yearly internal knots. The pale blue dotted line represented hazard ratio from an unadjusted Cox proportional hazards model and solid blue line represented hazard ratio from a Cox proportional hazards model adjusted for baseline age, sex, smoking status, socioeconomic quintile, influenza vaccine exposure in three years preceding study entry, consultation rate at medical practice in two years preceding entry, and updated Charlson comorbidity score. Shaded area represents 95% confidence intervals. Red solid line represents median follow-up time.

### **9. SUPPLEMENTARY RESULTS: ABSOLUTE MEASURES OF SURVIVAL**

The absolute survival gain for time-to-first-event calculated by restricted mean survival time at three years was 11.2 days (95% CI 10.5, 11.9 days) for death and 13.4 days (95% CI 12.1, 14.8 days) for infection-associated hospitalisation. A comparison of mean cumulative hospitalisation events in live shingles vaccine exposed and unexposed demonstrated that at three years the average cumulative number of events per individual was 1.55 and 1.84 respectively, equivalent to 290 fewer all-cause hospitalisations per 1000 individuals at three years (95% CI 230, 351). Live shingles vaccine exposure was associated with 59 fewer infection-associated hospitalisations (cumulative mean events 0.17 and 0.24 in live shingles vaccine exposed and unexposed respectively (95% CI 55, 64)).

### **10. REFERENCES**

1.	Lewis JD, Bilker WB, Weinstein RB, Strom BL. The relationship between time since registration and measured incidence rates in the General Practice Research Database. Pharmacoepidemiol Drug Saf. 2005 Jul;14(7):443–51. doi:10.1002/pds.1115 PubMed PMID: 15898131.

2.	Torisson G, Rosenqvist M, Melander O, Resman F. Hospitalisations with infectious disease diagnoses in somatic healthcare between 1998 and 2019: A nationwide, register-based study in Swedish adults. The Lancet Regional Health – Europe. 2022 May 1;16. doi:10.1016/j.lanepe.2022.100343 PubMed PMID: 35360441.

3.	Quan H, Li B, Couris CM, Fushimi K, Graham P, Hider P, et al. Updating and validating the Charlson comorbidity index and score for risk adjustment in hospital discharge abstracts using data from 6 countries. Am J Epidemiol. 2011 Mar 15;173(6):676–82. doi:10.1093/aje/kwq433 PubMed PMID: 21330339.

4.	Booth HP, Prevost AT, Gulliford MC. Validity of smoking prevalence estimates from primary care electronic health records compared with national population survey data for England, 2007 to 2011. Pharmacoepidemiology and Drug Safety. 2013;22(12):1357–61. doi:10.1002/pds.3537

5.	Raleigh V. The King’s Fund [Internet]. 2022 [cited 2023 Sep 12]. What is happening to life expectancy in England? Available from: https://www.kingsfund.org.uk/publications/whats-happening-life-expectancy-england

6.	Inns T, Fleming KM, Iturriza-Gomara M, Hungerford D. Paediatric rotavirus vaccination, coeliac disease and type 1 diabetes in children: a population-based cohort study. BMC Medicine. 2021 Jun 29;19(1):147. doi:10.1186/s12916-021-02017-1

7.	Therneau T, Crowson C, Atkinson E. ResearchGate [Internet]. 2024 [cited 2025 Jul 24]. Using Time Dependent Covariates and Time Dependent Coefficients in the Cox Model. Available from: https://www.researchgate.net/publication/265041278_Using_Time_Dependent_Covariates_and_Time_Dependent_Coefficients_in_the_Cox_Model

8.	Bozdogan H. Model selection and Akaike’s Information Criterion (AIC): The general theory and its analytical extensions. Psychometrika. 1987 Sep 1;52(3):345–70. doi:10.1007/BF02294361

9.	McCaw ZR, Yin G, Wei LJ. Using the Restricted Mean Survival Time Difference as an Alternative to the Hazard Ratio for Analyzing Clinical Cardiovascular Studies. Circulation. 2019 Oct 22;140(17):1366–8. doi:10.1161/CIRCULATIONAHA.119.040680

10.	Dong H, Robison LL, Leisenring WM, Martin LJ, Armstrong GT, Yasui Y. Estimating the Burden of Recurrent Events in the Presence of Competing Risks: The Method of Mean Cumulative Count. Am J Epidemiol. 2015 Apr 1;181(7):532–40. doi:10.1093/aje/kwu289 PubMed PMID: 25693770; PubMed Central PMCID: PMC4371763.
